# Spectral Analysis Comparison of Pushbroom and Snapshot Hyperspectral Cameras for *In-Vivo* Brain Tissues and Chromophores Identification

**DOI:** 10.1101/2024.06.06.24308500

**Authors:** Alberto Martín-Pérez, Alejandro Martinez de Ternero, Alfonso Lagares, Eduardo Juarez, César Sanz

## Abstract

**Significance:** Hyperspectral imaging sensors have rapidly advanced, aiding in tumor diagnostics for *in-vivo* brain tumors. Linescan cameras effectively distinguish between pathological and healthy tissue, while snapshot cameras offer a potential alternative to reduce acquisition time.

**Aim:** Our research compares linescan and snapshot hyperspectral cameras for *in-vivo* brain tissues and chromophores identification.

**Approach:** We compared a linescan pushbroom camera and a snapshot camera using images from 10 patients with various pathologies. Objective comparisons were made using unnormalized and normalized data for healthy and pathological tissues. We utilized Interquartile Range (IQR) for the Spectral Angle Mapping (SAM), the Goodness-of-Fit Coefficient (GFC), and the Root Mean Square Error (RMSE) within the 659.95 to 951.42 nm range. Additionally, we assessed the ability of both cameras to capture tissue chromophores by analyzing absorbance from reflectance information.

**Results:** The SAM metric indicates reduced dispersion and high similarity between cameras for pathological samples, with a 9.68% IQR for normalized data compared to 2.38% for unnormalized data. This pattern is consistent across GFC and RMSE metrics, regardless of tissue type. Moreover, both cameras could identify absorption peaks of certain chromophores. For instance, using the absorbance measurements of the linescan camera we obtained SAM values below 0.235 for four peaks, regardless of the tissue and type of data under inspection. These peaks are: one for cytochrome b in its oxidised form at *λ* = 422 nm, two for HbO2 at *λ* = 542 nm and *λ* = 576 nm, and one for water at *λ* = 976 nm.

**Conclusion:** The spectral signatures of the cameras show more similarity with unnormalized data, likely due to snapshot sensor noise, resulting in noisier signatures post-normalization. Comparisons in this study suggest that snapshot cameras might be viable alternatives to linescan cameras for real-time brain tissues identification.

## 1 Introduction

Cancer remains one of the leading causes of morbidity and mortality in the world, with approximately 18.1 million new cases of cancer diagnosed worldwide in 2020. In addition, the International Agency for Research on Cancer estimates that by 2040 the diagnosis of new cases will increase to 27.0 million.^1^ In Spain, 1.49% of newly diagnosed cancers in 2022 (4.169 of 280.101) were tumors of the encephalo or nervous system.^1^ Identifying pathological tissue from healthy tissue is challenging, especially with aggressive tumors such as grade IV glioblastoma (GB) that have high infiltration capabilities.^2^ Additionally, GB have poor long-term survival rates,^3^ making surgery an unavoidable process to increase patient survival. However, the brain shifting towards the skull opening can result in cerebrospinal fluid leakage and hinder tumor identification due to the alterations in the structure of the surrounding tissue, rendering pre-operative imaging inadequate for intra-operative conditions.^4^ Therefore, intraoperative tools for brain tumor surgery are essential for neurosurgeons. Existing tools such as neuronavigators, intraoperative magnetic resonances, intraoperative ultrasounds or add-on agents like 5-aminolevunilic acid (5-ALA)^5^ can help in delineating and locating the tumor, but each method also has limitations in pinpointing its exact location,^6^ increasing surgery time,^7, 8^ producing low-resolution images^9, 10^ and being invasive because of requiring the injection of an agent into the patient with a limited ability to define the tumor margin during surgery for low-grade gliomas.^11^ Therefore, faster and non-invasive techniques, compared to the tools described previously, are crucial to the success of surgical interventions. A widely used non-invasive and non-ionizing technique that requires no contact with patients is Hyperspectral (HS) Imaging (HSI).^12^ The advancement in HS sensors in the past years and the variety of available options can difficult the decision of which HS camera to use. In particular, HS cameras are capable of capturing spatial and spectral data using various techniques,^13^ being scanning-based (SB) and wide-field (WF) the typical imaging methods for HSI. Firstly, SB approaches can acquire the spectrum for each pixel using whiskbroom (point-scanning) instruments, a line of pixels in pushbroom (line-scanning) instruments, or by using a wedge filter that disperses light spectrally along one dimension (wedge-scanning). Secondly, WF approaches capture the whole scene in a single exposure with 2-D detector arrays, either by stepping through the wavelength spectrum to complete the data cube (wavelength scan) or by acquiring the spatial and spectral information at the same time (snapshot). However, recent techniques such as snapscan cameras,^14^ which combine SB and WF approaches, provide compact solutions with faster acquisition times than linescan cameras while offering higher spatial and spectral resolution than snapshot sensors. It is worth noting that the most used spectral range in medical applications fall in the Visible (VIS) (400 to 780 nm) and Near-Infrared (NIR) spectrums (780 to 2.500 nm).^12^ Regardless of the possibilities available to select HSI equipment, pushbroom linescan, snapscan and snapshot HS cameras have been used as part of intraoperative tools in several studies to differentiate brain tumor from healthy tissue in *in-vivo* human brains.^15–18^ For example, Fabelo et. al. developed an intraoperative acquisition system based on two pusbroom linescan cameras, a VNIR and a NIR in the spectral range between 400 to 1.000 nm and 900 to 1700 nm, respectively.^15^ The intraoperative acquisition and data processing took approximately 1 minute, which does not enable for real-time solutions understood as providing a live sequence of images. Furthermore, other works by Vandebriel et. al. have assembled a snapscan HS camera to a surgical microscope for improving the removal of low grade gliomas.^16^ The camera captured 104 spectral bands between 470 to 787 nm since it had to match the intrinsic illumination of the microscope. Even though the snapscan camera can provide high spatial resolution in a wide spectral range and reduce the acquisition time to less than 3 seconds for static targets,^14^ it requires an internal movement of a linescan sensor to acquire a HS cube, which is not suitable for real-time solutions. Moreover, recent works have developed an intraoperative system based on HSI with a pushbroom linescan and a snapshot HS cameras^18, 19^ for brain tumor detection. On one hand, the snapshot camera captures 25 spectral bands in the 660 to 950 nm spectral range with a spatial dimension of 409×217 pixels each band. On the other hand, the pushbroom linescan camera acquires 369 bands between 400 to 1.000 nm with a spatial dimension of 1600 pixels each line. Despite the low number of bands acquired by the snapshot camera and its low spatial resolution, it can enable for real-time solutions such as live video classifications with machine learning algorithms.^19^

Therefore, this research study aims to compare two different HS cameras that differ on how they acquire data, a pushbroom linescan and a snapshot, to determine their potential use for brain tissues and chromophores identification. To compare how similar their signatures are, the analysis is conducted in the NIR spectrum, specifically in the 659.95 to 951.42 nm range. The study involved examining and using ten different *in-vivo* human brain images from the Slim Brain database^20^ to provide useful results for biomedical purposes. All the brains from patients used from the database were captured with both HS cameras to ensure similar lighting conditions for the comparisons. Sec. 2 describes the cameras and acquisition system used in the study, the images used, how those are processed as well as the spectral similarity metrics to compare the measurements taken with both HS cameras. Next, Sec. 3 presents the results while Sec. 4 provides a discussion with the findings of the study. Finally, Sec. 5 gives the conclusions of this study.

## 2 Materials and Methods

### 2.1 Hyperspectral Camera Specifications

Two different HS cameras based on different acquisition techniques have been used to capture *in-vivo* brain images. The technical specifications of both cameras, sensors and lenses are presented on Table 1.

**Table 1.**
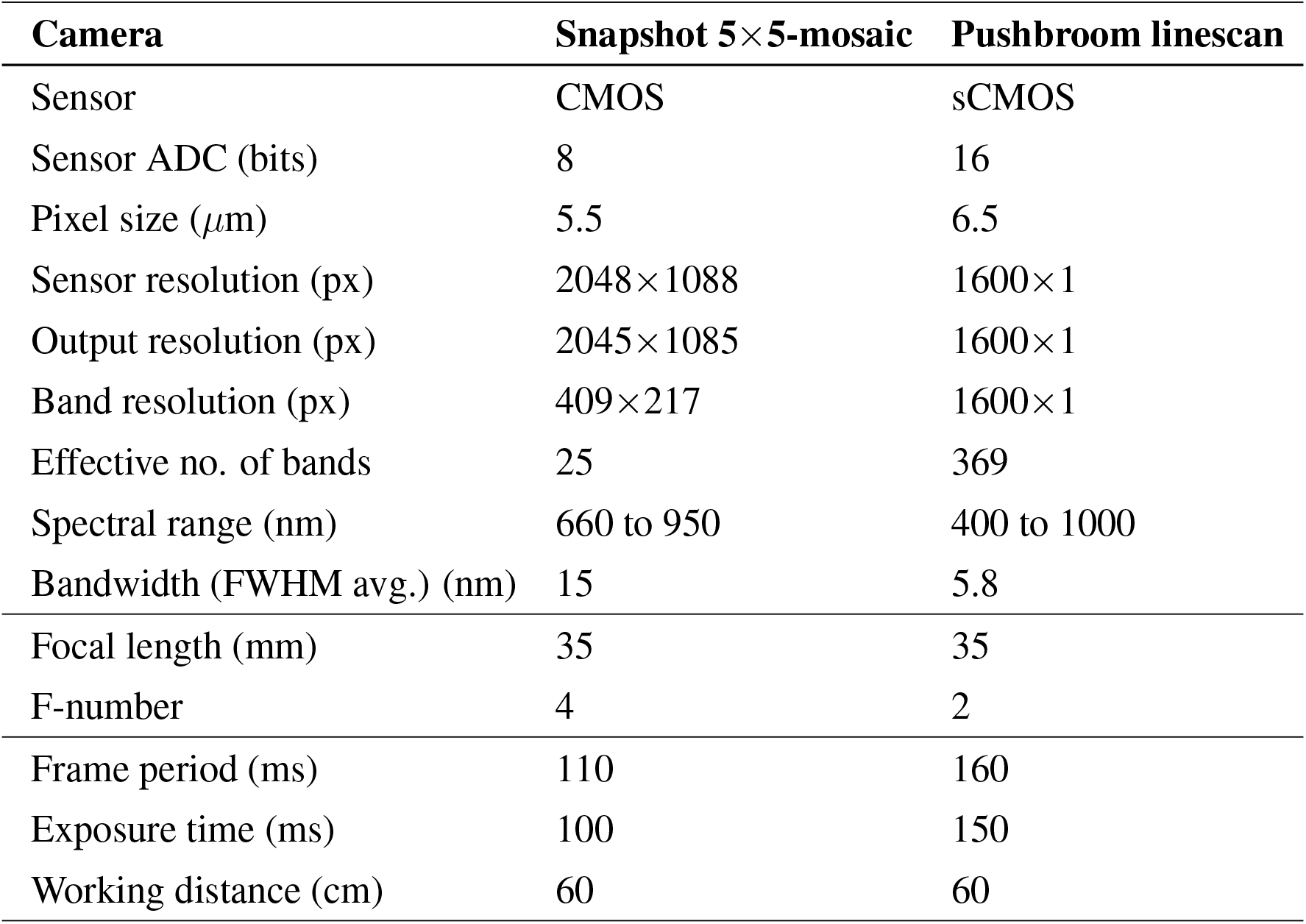
Sensor and camera optics specifications of the different hyperspectral cameras used. Last five parameters are fixed during the acquisition of images at the operating room.

On one hand, the snapshot HS camera has a complementary metal oxide semiconductor (CMOS) sensor holding a 5×5-mosaic pattern with a pixel size of 5.5 *μ*m (MQ022HG-IM-SM5X5-NIR, Ximea GmbH, Germany). The analogue to digital converter (ADC) of the sensor provides images with a resolution of 8 bits. Additionally, a long pass filter (FELH0650, Thorlabs, Inc., USA) with a 650 nm cut-on wavelength is placed in front of the lens to remove non-negligible secondary harmonics, which were determined by the manufacturer in the spectral response curves during sensor production. Although the sensor resolution is 2048×1088 pixels, the active area of the sensor with the built-in spectral filters has 2045×1085 pixels. This reduced area is called the active filter zone, which omits the last 3 rows and last 3 columns from the total sensor resolution. Each capture with this snapshot camera produces an image containing the spatial and spectral resolution due to the 5×5-mosaic pattern. Each mosaic contains approximately the same spatial pixel at different 25 wavelength bands within the NIR spectrum, specifically in the 660 to 950 nm spectral range. Additionally, these bands are spaced among each other with a mean and standard deviation value of 12.11 ± 2.64 nm. Hence, the mosaic pattern reduces the two-dimensional output spatial resolution by a factor of 5 to obtain a three-dimensional HS cube. In particular, the image with 2045×1085 pixels generated by the sensor is arranged into a 409×217×25 HS cube to perform the spectral analysis. The main advantage of the snapshot camera is its capability for real-time solutions, understood as processing a sequence of HS images to provide a live video of the scene. On the other hand, the other camera is based on the pushbroom linescan technology (Micro-Hyperspec® E-Series, HeadWall Photonics Inc., USA), which holds a scientific CMOS sensor with an ADC of 16 bits and a pixel size of 6.5 *μ*m. The sensor acquires a single spatial line with 1600 pixels and 394 wavelengths of information. Thus, the camera needs to be moved with an actuator to scan an image with as many lines as desired. All *in-vivo* brain captures used in this study were scanned with 500 lines, producing images with a spatial resolution of 1600×500 pixels and 394 spectral bands. Besides, the exposure time and frame period were set to 150 ms and 160 ms, respectively. Although the sensor is sensitive in the 365 to 1004 nm spectral range, those wavelengths acquired outside the 400 to 1000 nm spectral range need to be removed, as specified by the manufacturer. Eliminating such bands results in 369 effective wavelength bands separated by 1.62 ± 0.00 nm from each other.

It is worth noting that captures from both cameras were cropped spatially to help neurosurgeons during the labelling processing. Therefore, the spatial resolution of the HS linescan or snapshot captures are smaller than 1600×500 pixels or 409×217 pixels, respectively. Although the pusbroom linescan camera provides more spatial resolution and spectral information, it is not suitable for real-time solutions due to the scanning procedure. The time spent to scan a brain image requires, approximately, 1 minute and 40 seconds, while an image captured with the HS snapshot camera takes 100 ms.

### 2.2 Acquisition System

The acquisition system used to gather the *in-vivo* brain images is presented in Fig. 1. Starting from the left, the HS 5×5-mosaic snapshot camera is located inside a 3D printed white case. Inside the case, there is a servomotor employed to help focusing the camera. An external light source with a 150 W 21 V EKE halogen lamp (MI150, Dolan-Jenner, USA) and a dual gooseneck fiber optic (EEG28, Dolan-Jenner, USA) were used to illuminate the brains. While the housing of the light source does not appear in the image, the dual fiber optics are shown glowing in Fig. 1. For this study, the laser imaging detection and ranging (LiDAR) is used to measure the working distance of the acquisition system regarding the brain surface. Given that the linescan camera has a fixed focusing distance of 60 cm, the measurement of the LiDAR is necessary to ensure focused captures with the linescan camera. Therefore, all images were taken at a distance of 60 cm to ensure a fair comparison between the two cameras, taking into account the limited focus distance of the linescan camera. Furthermore, the motorized linear stage (X-LRQ300HL-DE51, Zaber Technologies Inc., Canada) below the imaging sensors is mainly used to move the HS linescan camera. This movement allows the scanning of the brain image to compose a HS cube with the desired spatial resolution. The sensors and fiber optics are all aligned in the same plane, which is perpendicular to that of the linear stage holding them. Although not included in Fig. 1, two additional motorized linear stages are used to tilt and provide height to the stage.

**Fig 1.**
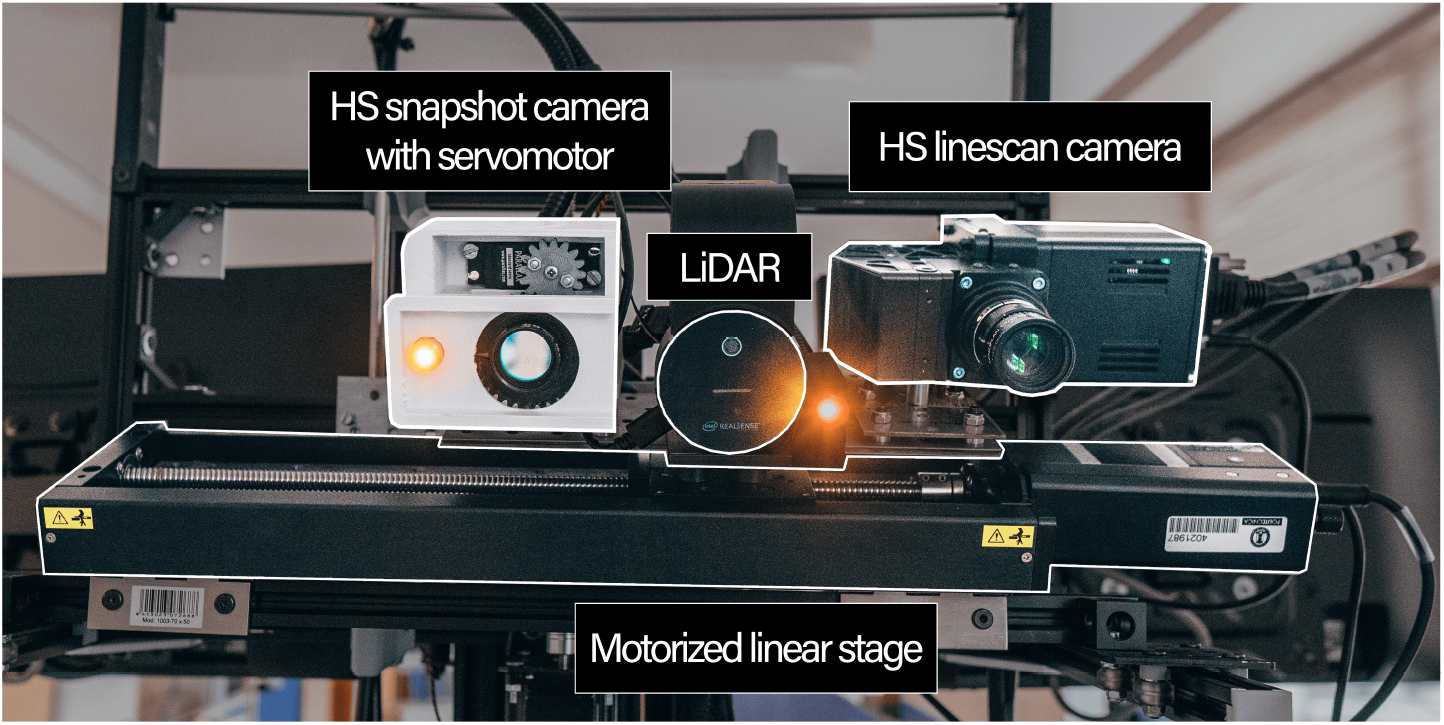
Front view of real acquisition system with main components highlighted and labeled.

### 2.3 Data Processing

All HS images, regardless of the camera used, are pre-processed using almost the same procedures, which are presented in Table 2.

**Table 2.** Pre-processing steps performed to the images captured with the different HS cameras used.

| Camera | Snapshot $5 \times 5$-mosaic | Pushbroom linescan |
| --- | --- | --- |
| 1. Radiation calibration | ✓ | ✓ |
| 2. Cube formation | ✓(Rearrange of 2D image) | ✓(Line stitching) |
| 3. Spectral correction | ✓ | ✗ |
| 4. Bands removal | ✗ | ✓(Extract effective bands) |
| 5. Noise filtering<sup>21</sup> | ✓ | ✓ |
| 6. Normalization | ✓ | ✓ |

The first step is to obtain the reflectance information of the material interrogated by calibrating the raw information obtained with the sensor. Eq. 1 eliminates the effect of the HS sensor and the lighting conditions captured with the raw images to obtain the reflectance information *R* from the sample,

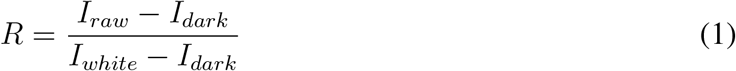

where *I*_*raw*_ is the captured raw data of the sample, *I*_*dark*_ is the raw dark reference captured with the lens cap in front of the camera lens, and *I*_*white*_ is the raw white reference intensity reflected over a Lambertian diffuse target with 95% of reflectance values (SG 3151-U, SphereOptics GmbH, Germany). All white references are captured under the same conditions as the images captured, meaning that each calibrated image needs a *I*_*white*_ captured with the same working distance and tilt angle. While *I*_*dark*_ is used to remove the ambient temperature and electrical noise introduced to the measurement, *I*_*white*_ tries to reduce the influence of the light sources on the sample. The second step is related to the formation of the HS cubes. The snapshot sensor captures 2D images which need to be rearranged into a 3D cube, which has a spatial dimension 5 times smaller than the 2D images due to the mosaic pattern of the sensor described in Subsec. 2.1. However, the pushbroom camera have a simpler process to conform a HS cubes. Such process is called line stitching, which requires a precise linear actuator to move the HS camera to join adjacent spatial lines while avoiding overlap. The spectral correction is the third step and only applies to the snapshot camera. This correction is necessary to correct the response curves of the HS snapshot sensor, which presents crosstalks between adjacent pixels of the sensor that vary with the angle of incident light to the sensor.^22^ To correct this effect, the manufacturer provides a spectral correction matrix that modifies the response of the sensor to obtain ideal Gaussian curves. The spectral correction process is described in Eq. 2:

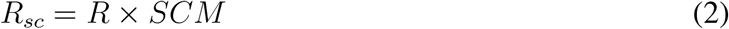

where *R*_*sc*_ is the spectrally corrected reflectance data, *R* is the reflectance of the sample after using Eq. 1 and *SCM* is the spectral correction matrix. The forth step is the removal of bands, which only applies to the pushbroom camera. As described in Subsec. 2.1, the sensor captures more information than what can be effectively used. Briefly, this process eliminates the spectral bands not taken between the 400 to 1000 nm spectral range. The fifth step consists on applying the noise filtering algorithm of HySime, which was presented by Bioucas-Dias et. al.^21^ We specifically use the noise estimation procedure of HySime and assume that the noise in the HS cube is additive. In such procedure, HySime deduces the noise present in a HS cube by making the assumption that the reflectance at a specific band can be effectively characterized through linear regression using the remaining bands. After estimating the noise in a HS cube, we employ this estimation to subtract the noise from the original HS cube, thus carrying out the noise filtering process for each image independently. The last step is a normalization process which homogenize the spectral signatures to help comparing captures. For this study, the normalized reflectance *R*_*norm*_ is obtained using a min-max normalization for every spectral pixel independently, as described in Eq. 3, which forces data to a range of 0 to 1:

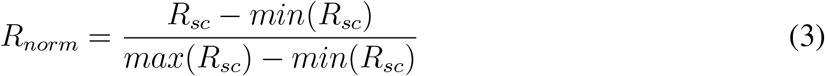

where *R*_*sc*_ is the spectrally corrected reflectance data obtained with Eq. 2 for the HS snapshot camera or the reflectance information *R* obtained with Eq. 1 for the HS linescan camera. For further clarification, in Eq. 3 the noise filtering is applied to either *R*_*sc*_ or *R* before applying the normalization.

### 2.4 In-vivo Human Brain Images

For this study we used ten *in-vivo* brains from adult patients who have provided an informed consent prior to surgery. These ten patients were selected because two or more sterilized rubber rings were placed by neurosurgeons prior to acquire the HS images. Different colours were used for tissue identification, with green and black being used for healthy and pathological tissues, respectively. These rubber rings can be seen in the pseudo RGBs of Fig. 2 for patient with ID 190. Additionally, the same images for the rest of the patients are presented in Fig. S1 of the Supplementary Materials. As indicated previously, images of patients who have suffered different pathologies have been used. More specifically, patients with ID 177, 183, 184, 190, 193, and 203, have GB with Isocitrate Dehydrogenase (IDH) non-mutated. Patient ID 185 has grade II astrocytoma with IDH mutation, patient ID 192 has metastatic testicular tumor, and patient with ID 194 has metastatic lung carcinoma tumor. Last but not least, patient ID 201 exhibits pseudoprogression in a GB with IDH non-mutated, indicating apparent GB progression likely attributable to treatment effects rather than actual tumor growth.

**Fig 2.**
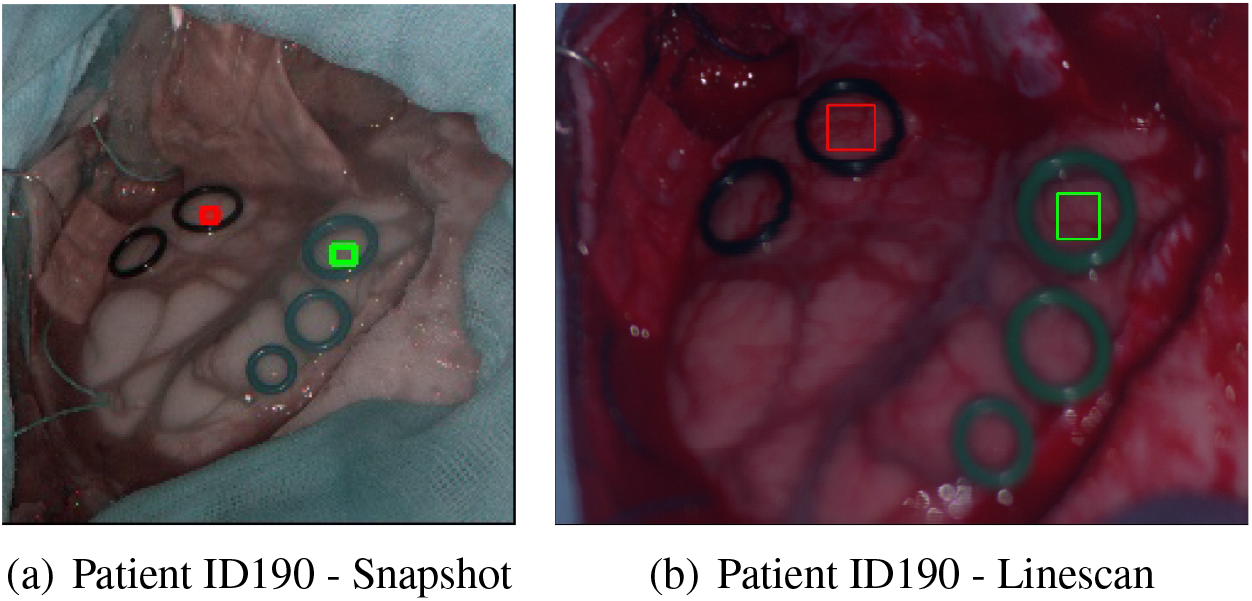
Pseudo RGB with healthy and pathological region of interests (ROIs) for patient ID 190.

The guidelines of the Declaration of Helsinki have been followed and the acquisition of HS images have been approved by the Research Ethics Committee of Hospital Universitario 12 de Octubre, Madrid, Spain (protocol code 19/158, 28 May 2019). All patients shaved the area to be operated on prior to the scalp incision. Then, a high-speed drill was employed to make burr holes in the skull, which are used to insert a cranial drill to perform the craniotomy. This procedure extracts a bone flap to expose the dura of the patient, and then the durotomy is performed by cutting with a knife the dura to uncover the brain surface. Then, both HS cameras proceeded to acquire the *in-vivo* brain surface.

Using the rubber rings ensures that the data of both cameras comes from the same spatial location of the brain surface. Furthermore, this procedure is similar to the one employed by Mühle et. al.,^23^ who used a plastic cursor to delimit a bordered region of interest (ROI) over biological organs to compare different HS cameras. In addition, the biomedical experts selected similar areas of size 21×21 pixels inside the plastic cursor to perform their analysis. In our case, the areas we selected inside the rubber rings have a size of 5×5 pixels since bigger ROIs got pixels outside the rings in the snapshot images. Although in the previous study Mühle et. al. took ten HS snapshot images to average them, in our study it was not feasible sTince the *in-vivo* human brain is in motion due to the heartbeat. Moreover, the linescan camera is inevitably affected by the motion of the brain during the scanning procedure, hindering the possibility to average multiple captures. For these reasons, one HS snapshot and one HS linescan image were taken for each patient. A single linescan image takes approximately 1 minute and 40 seconds, while a snapshot measurement takes 100 ms. Notice that specular reflections have not been removed, as neither the cameras nor the light source had polarizers. Although the patients exhibit diverse pathologies, the spectral comparisons are conducted on an intra-patient basis. This entails the comparison of the spectral signatures of both cameras for each patient individually.

### 2.5 Spectral Similarity Metrics

A way to compare both HS cameras is to employ spectral similarity metrics, which can asses how different reflectances are related to each other. Agarla et. al. have examined 14 frequently used measures and grouped them into five categories based on the type of error they evaluate,^24^ including the mathematical definition and implementation of all metrics. Furthermore, as stated by Agarla et. al., selecting one measure for each of the groups they describe can be sufficient to asses the spectral similarity.^24^ For that reason, we decided to use the Root Mean Square Error (RMSE), the Goodness-of-Fit Coefficient (GFC),^25^ and the Spectral Angle Mapper (SAM)^26^ metrics. On one hand, RMSE can range from 0 to 1 since the maximum value of our data is 1, indicating a value of RMSE equal to 0 perfect similarity. On the other hand, GFC values can be between the 0 to 1 range, where a value of 1 indicates complete similarity. SAM values range from 0 to 1, indicating values closer to 0 high similarity and values closer to 1 low similarity.

These metrics are chosen since they are applicable to the spectral domain, can be used as loss functions (except SAM since AM are not used to measure losses) and do not need extra requirements to be computed. Furthermore, the spectral signatures of the HS linescan camera are considered as the reference spectra, while those obtained with the HS snapshot camera are considered as the arbitrary signal. The previous assumption is grounded on the fact that the HS linescan camera gathers greater spatial and spectral data compared to the snapshot camera. We compute and compare the mean spectral signatures as illustrated in Fig. 3. For each patient, we independently obtain the mean spectral signatures of the pixels in the ROIs located inside the rubber rings, as shown in the left side of Fig. 3.

**Fig 3.**
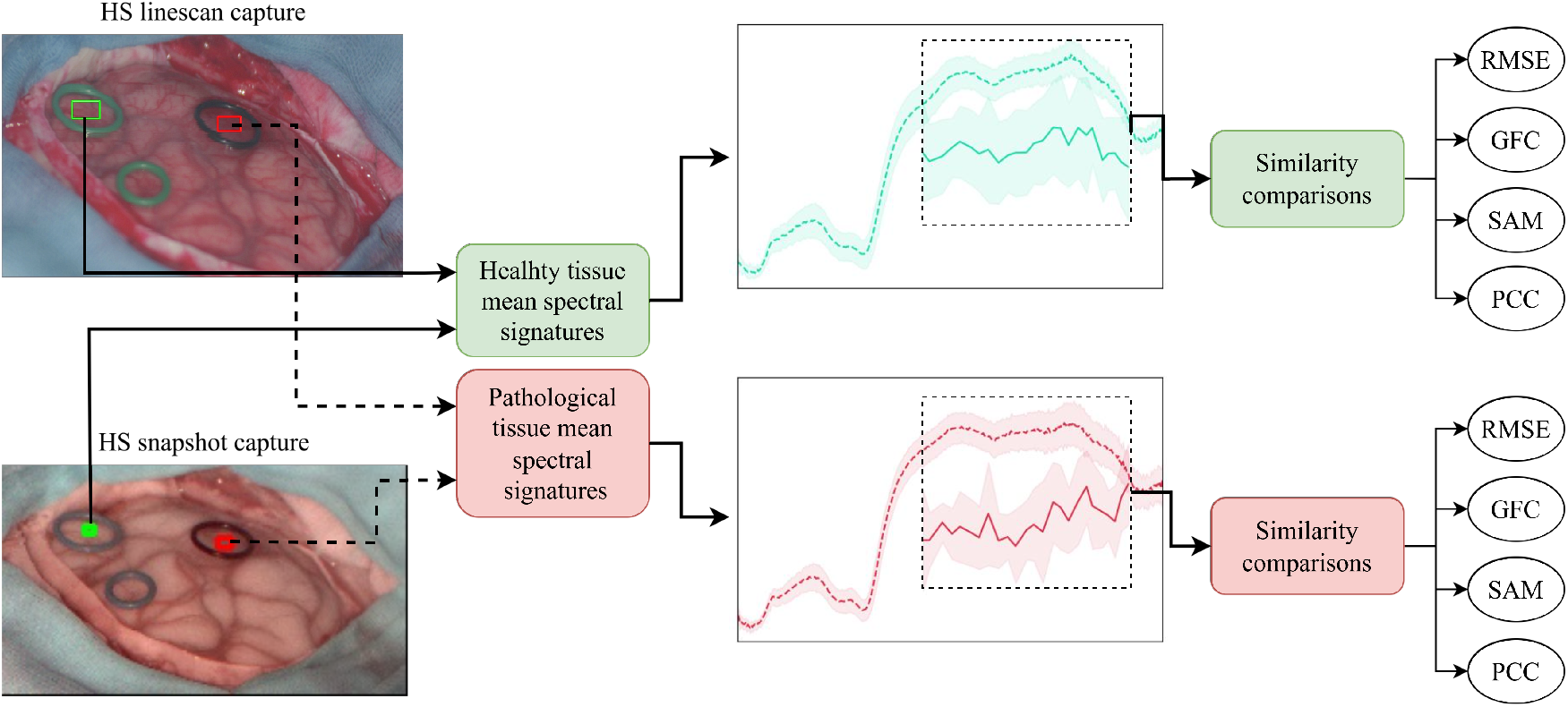
Procedure to compute the spectral similarities acquired with both HS cameras. The comparison is made independently by patient and tissue, using the spectra shared by both cameras.

Since the HS snapshot camera captures less spatial resolution, less pixels are included in the green and red ROIs inside the rubber rings compared to those captured by the HS linescan. Specifically, the ROIs created for all snapshot captures have a 5×5 spatial dimension. Therefore, to provide a fair comparison we created bigger ROIs for the HS linescan but randomly selected 25 pixels. Then, with those 25 pixels for each tissue and camera, the mean spectral signature and standard deviation are computed as presented with the plots in the middle of Fig. 3, which include black dashed rectangles to indicate the part of the spectrum shared by both cameras and used to compare them with the spectral similarity metrics. Lastly, the *RMSE, GFC*, and *SAM* metrics are computed for each tissue using the matched wavelengths of the cameras presented in Table S1, available in the supplementary materials.

### 2.6 Analyzing Absorbance Measurements to Identify Chromophores Absorption peaks

To analyze the absorbance (*A*) spectral signatures and look for patterns that might indicate the presence of certain chromophores, we will use the reflectance (*R*) measured by the cameras. *A* is commonly derived,^27–29^ for each wavelength, from *R* using Eq. 4:

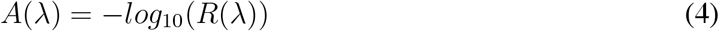

This expression is derived from the Beer-Lambert law,^30^ which expresses *A* as *A* = log_10_(*I*_*o*_*/I*). Here, *I*_*o*_ represents the incident light, and *I* is the light that has passed through the sample. In our specific context, we are dealing with reflectance information as defined in Eq. 1. The maximum reflected light corresponds to *I*_*white*_ (analogous to what would be *I*_*o*_ in the Beer-Lambert law), and *I*_*raw*_ represents the light that has passed through the brain (similar to *I* in the Beer-Lambert law). To account for the noise of the sensor, we include the dark measure from Eq. 1 (*I*_*dark*_). This allows us to mitigate sensor noise, and as a result, we arrive to Eq 4: *A* = log_10_((*I*_*white*_ − *I*_*dark*_)*/*(*I*_*raw*_ − *I*_*dark*_)) = log_10_(1*/R*) = − log_10_(*R*).

The chromophores we will attempt to identify are those that contribute the most to absorb light in the NIR region (from 800 to 2,500 nm) in adult brains. As stated by Correia et. al. these are hemoglobin, water, lipid and the following cytochromes: cytochrome aa3 (Cyt aa3), cytochrome b (Cyt b), and cytochrome c (Cyt c).^31^ The absorption spectra of the previous cytochromes on their redox state between 400 and 1,000 nm were obtained from the Biomedical Optics Research Laboratory (BORL) Github repository.^32^ It is worth noting that we converted the molar extinction coefficient for the Hb to absorption coefficient as specified by Prahl. et. al.,^33^ considering that for whole blood there is 150 g of Hb per liter. Although this assumption may be doubtful for the measurements taken, it allows us to visually compare the spectrum of all chromophores with the same units. Also, it is worth noting that the absorbance, *A*, and the absorption coefficient, *μ*_*a*_, represent different phenomena. On one hand, *A* is a property of a material that measures the fraction of light that can pass through in terms of intensity. On the other hand, *μ*_*a*_ is a property of the material that describes its effectiveness in absorbing light. We know from the Beer-Lambert law^30^ that *A* and *μ*_*a*_ are related through Eq. 5:

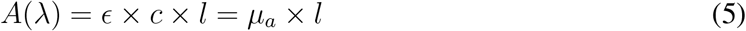

where *ϵ* is the molar absorptivity, also called the extinction coefficient, with units of 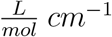, *c* is the concentration of a solution in the sample in 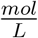, *l* is the length of the sample that light passes through in *cm*, and *μ*_*a*_ = *ϵ* × *c* is the absorption coefficient in *cm*^−1^. Unfortunately, *l* is unknown for the brain images used in this study. Therefore, we can not convert the *A* measured with the HS cameras to *μ*_*a*_, for comparing the chromophores spectra with the absorbance measurements of the cameras. However, we can use the SAM metric described in Subsec. 2.5 to try to identify the *μ*_*a*_ peaks of the chromophores in the *A* measurements. Of all metrics used in this study, SAM is the only one that focuses on the shape of the spectra rather than the numerical values.^24^ Although we cannot indicate the percentage concentration of each chromophore in the camera measurements, we try to identify their peaks by selecting wavelengths that include the *μ*_*a*_ peak wavelength and those around it. The most relevant absorption peaks of each chromophore are presented in Fig. X, available in the Supplementary Materials.

## 3 Results

### 3.1 Brain Tissue Measurements

The reflectance measurements obtained with both HS cameras from the 10 *in-vivo* human brains are shown in the Supplementary Materials, specifically in Fig. S2. The illustrated spectral signatures have been obtained, for every camera measurement, using 25 pixels located inside the rubber rings as shown in the images in Fig. 2. We decided to analyze the effect of normalizing the calibrated and denoised data to check how it would influence on the spectral similarity metrics. Hence, the two columns to the left in Fig. S2 are data that have been calibrated and denoised, while the two columns to the right are the same data which have been normalized using Eq. 3. The spectral range analyzed to address the comparison is between 659.95 to 951.42 nm using 25 bands detailed in Table S1 for each camera. To get an overview of the measurements, we illustrate in Fig. 4 the averaged results of all patients for both tissues presented in Fig. 4. The circular marks indicate the closest wavelengths between cameras specified in Table S1 available in the Supplementary Materials. By zooming into the common spectral range of the cameras between 659.95 and 951.42 nm from the previous plots, we present subfigures (c) and (d) to address in further detail the difference between measurements. Note the influence of the infrared peak emmitted by the LiDAR at *λ* ≈ 850 nm, illustrated in the dashed rectangles. Such peak is only present in the snapshot measurements since the LiDAR could only be turned off when capturing with the linescan. By normalizing the pixels from subfigures (c) and (d) we obtained the mean spectral signatures with standard deviation for both tissues in subfigures (e) and (f).

**Fig 4.**
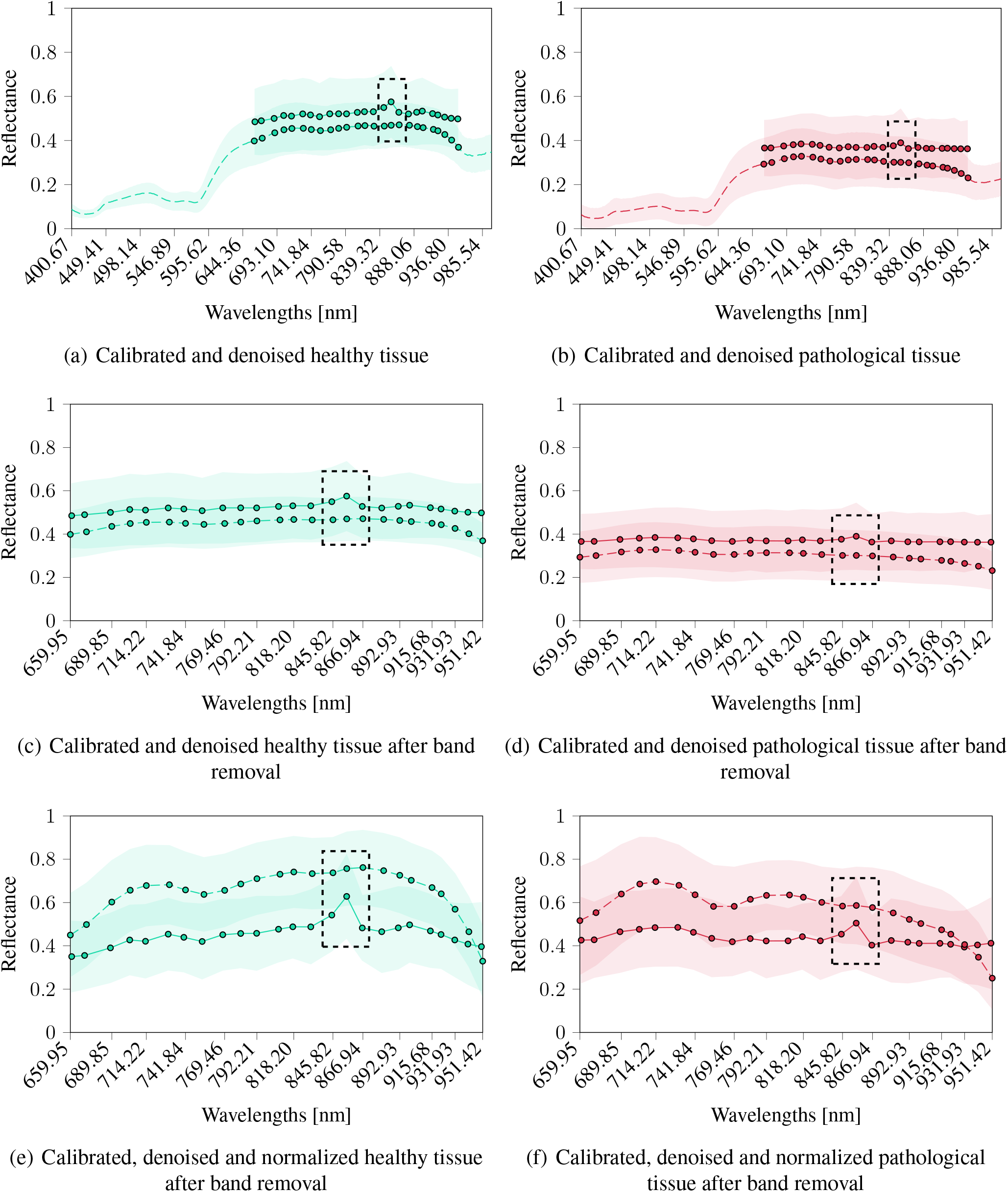
Mean reflectance spectral signatures with standard deviation of all pixels included inside the 5×5 ROIs from both HS cameras. The shorter spectral signatures from 661.61 to 951.42 nm nm with continuous lines correspond to the snapshot camera, while the longer spectral signature with dashed lines correspond to the linescan camera. The dashed rectangles indicate the spectral bands of the HS snapshot camera influenced by the infrared of the depth camera.

The analysis comparing the absorbance measurements using both HS cameras with the absorption coefficient spectra of deoxy-hemoglobin (Hb), oxy-hemoglobin (HbO2),^33^ Cyt aa3, Cyt b, Cyt c,^32^ and mammalian fat^34^ is illustrated in Fig. 5 and Fig. 6. In addition, a detailed analysis of the results obtained with respect to the absorbance measurements is provided in Section 4 of the Supplementary Materials, including Fig. S3 with all relevant absorption peaks for the chromophores under analysis.

**Fig 5.**
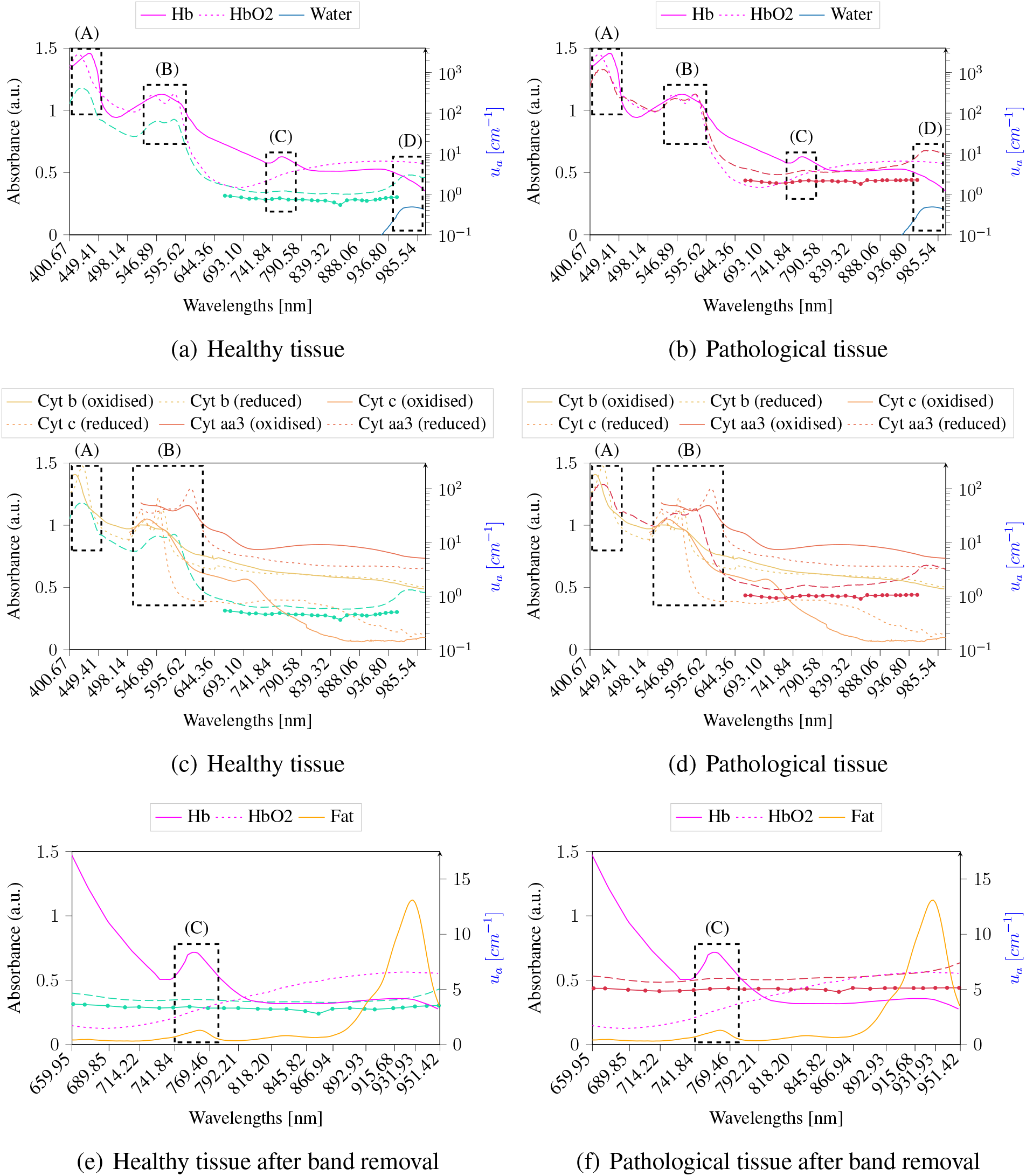
Mean absorbance spectral signatures of the 25 pixels included inside the rubber rings for both HS cameras. Data is calibrated and denoised. The shorter spectral signatures from 659.95 to 950.64 nm with continuous lines correspond to the snapshot camera, while the longer spectral signature with dashed lines correspond to the linescan camera. The spectra of Hb and HbO2^33^ are shown in (a) and (b) with continuous and dashed magenta lines, respectively, whereas cytochromes b, c, and c aa3^32^ are shown in (c) and (d) with different shades of oranges. Furthermore, continuous orange lines correspond to the absorption coefficients spectra of mammalian fat.^34^ Hb, HbO2, Cyt. b, Cyt. c, Cyt. c aa3, and fat are in *cm*^−1^, whose scale is in the right y-axis, whereas absorbance data measured with both cameras have its scale in the left y-axis. The A, B, and C dashed rectangles are used to indicate different absorption peaks of Hb, HbO2, Cyt. b, Cyt. c, Cyt. c aa3, or fat, while the D rectangle points out a water absorption peak at *λ* = 976 nm.

**Fig 6.**
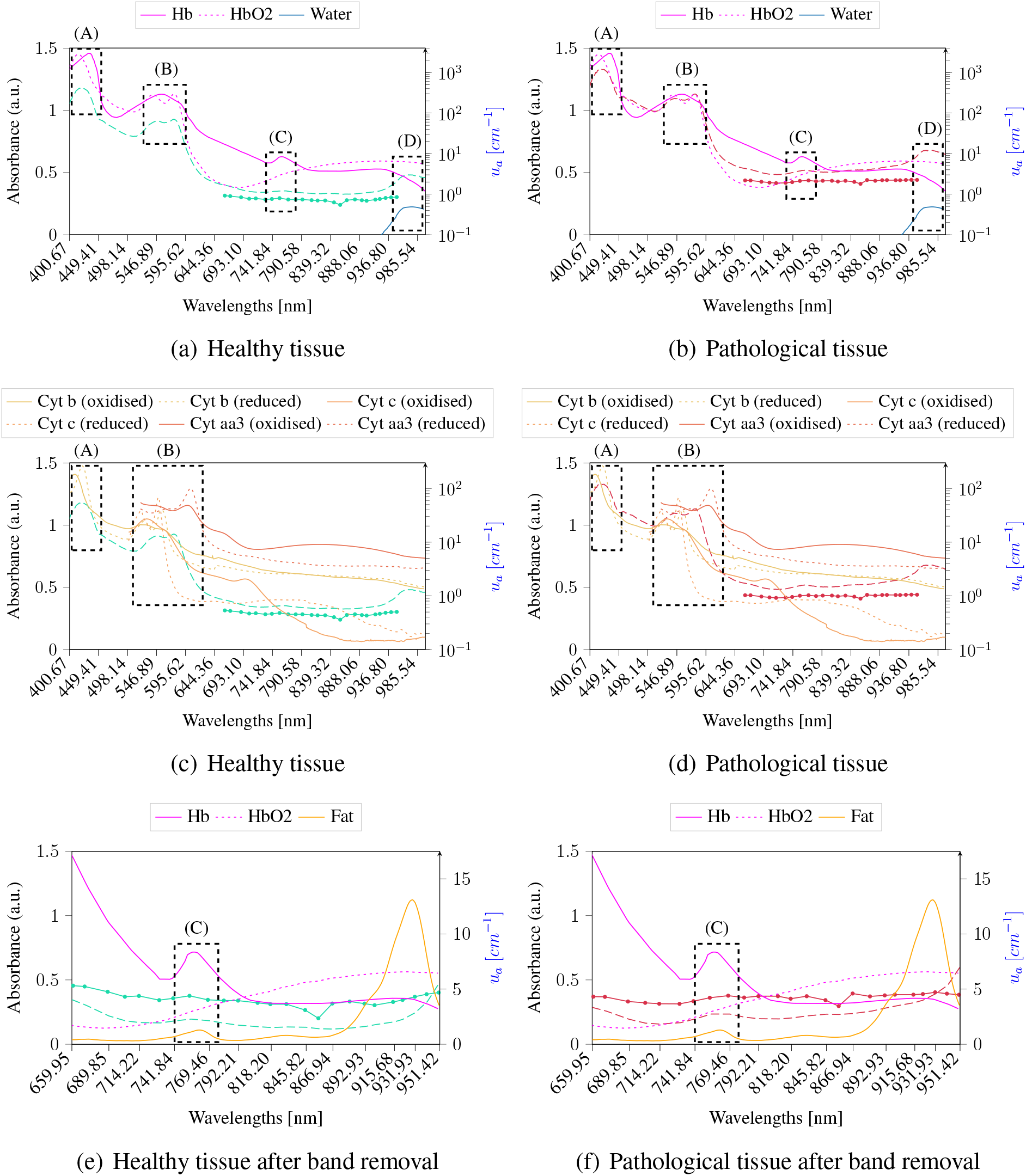
Mean absorbance spectral signatures of the 25 pixels included inside the rubber rings for both HS cameras. Data is calibrated, denoised and normalized. The shorter spectral signatures from 659.95 to 950.64 nm with continuous lines correspond to the snapshot camera, while the longer spectral signature with dashed lines correspond to the linescan camera. The spectra of Hb and HbO2^33^ are shown in (a) and (b) with continuous and dashed magenta lines, respectively, whereas cytochromes b, c, and c aa3^32^ are shown in (c) and (d) with different shades of oranges. Furthermore, continuous orange lines correspond to the absorption coefficients spectra of mammalian fat.^34^ Hb, HbO2, Cyt. b, Cyt. c, Cyt. c aa3, and fat are in *cm*^−1^, whose scale is in the right y-axis, whereas absorbance data measured with both cameras have its scale in the left y-axis. The A, B, and C dashed rectangles are used to indicate different absorption peaks of Hb, HbO2, Cyt. b, Cyt. c, Cyt. c aa3, or fat, while the D rectangle points out a water absorption peak at *λ* = 976 nm.

### 3.2 Reflectance Spectral Similarities to Compare Both Cameras

The spectral similarities between the reflectances measured with both HS cameras are presented in Fig. 7. This figure shows raincloud plots^35^ for the healthy and pathological tissues with green and red colors, respectively. Raincloud plots are a useful graphical representation that addresses the challenge of data obfuscation in the presentation of error bars or box plots. These visualizations combine various data elements to display raw data points, probability density through half violin plots, and key summary statistics such as median, first and third quartiles, outliers with black diamonds, and relevant confidence intervals via boxplots. This combination produces a visually appealing and adaptable representation with minimal repetition. Additionally, these plots have a red dot inside each box plot to illustrate the mean value of each distribution.

**Fig 7.**
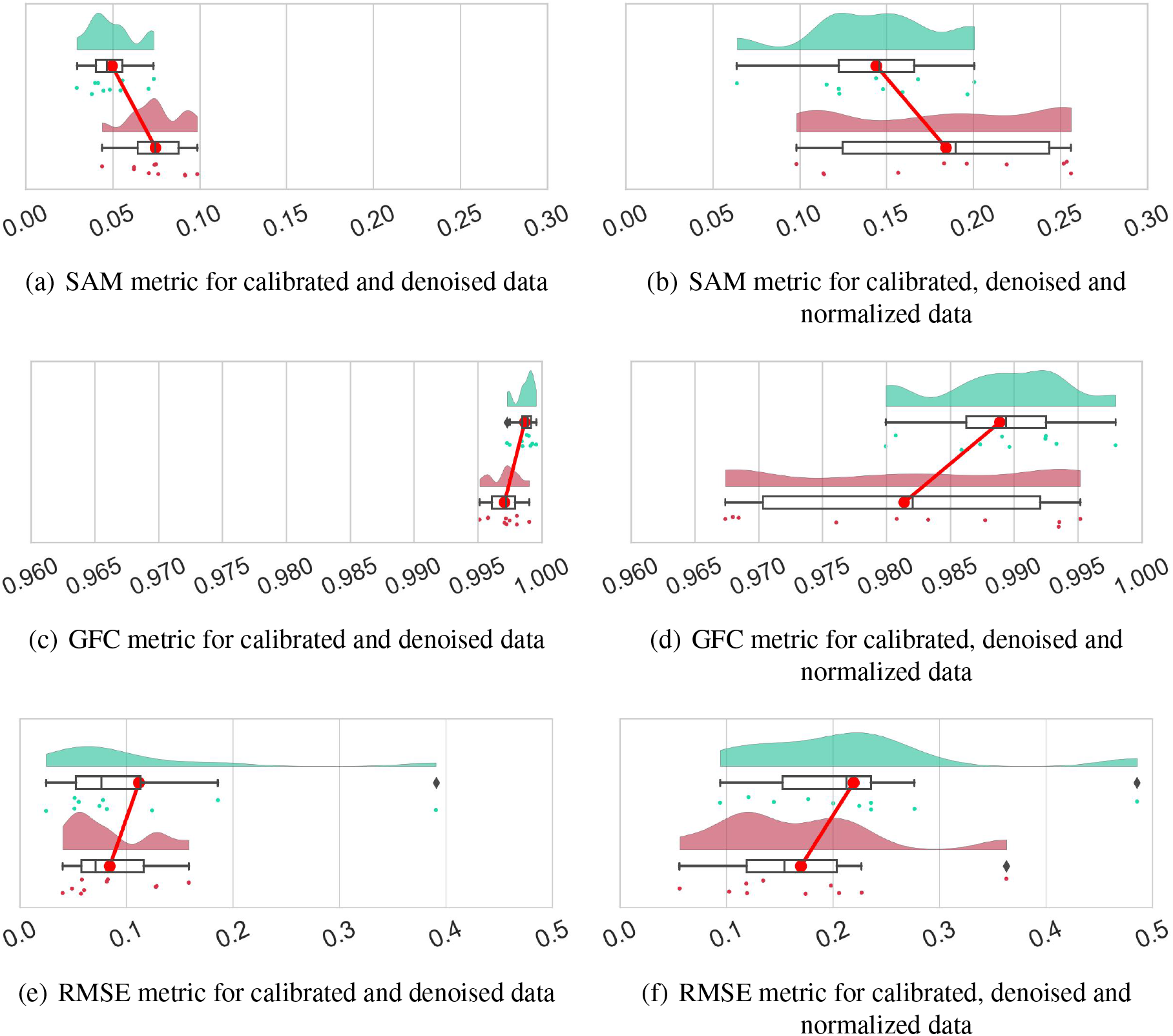
Raincloud plots with SAM, GFC, and RMSE similarity metrics. The left column plots ((a), (c), (e) and (g)) are computed when pixels in the ROIs have been calibrated and denoised, while plots in the right column ((b), (d), (f) and (h)) have additionally been normalized (after removing non-matched wavelengths of the HS linescan camera) using the min-max normalization from Eq. 3. Green distributions and dots indicate healthy tissue, while in red they indicate pathological tissue. There is 1 dot in every distribution for every patient described in Subsec. 2.4. Big red dots joined by a red line indicate the mean value of the distribution.

The pair of red dots shown in every plot of Fig. 7 are also connected to each other with a red line to visually see which distribution has a higher mean value. Furthermore, in these charts, each dot located below the box plots represents a single value of the corresponding similarity metric, which refers to the comparison of the mean spectral signatures for a particular patient and tissue type. For example, the leftmost green dot in Fig. 7 (a) represents the SAM value of 0.029 obtained when comparing the healthy tissue of both cameras for the patient with ID 193.

For comparison purposes, each row in Fig. 7 shows a pair of raincloud plots for every metric described in Subsec. 2.5. Plots located to the left of the figure are the similarity results computed when data were calibrated and denoised (Figs. 7 (a), (c), (e) and (g)), whereas those to the right are obtained when data were additionally normalized (Figs. 7 (b), (d), (f) and (h)). The obtained metrics for the comparison of each patient, including both tissues under analysis, are presented in Table S2 for further analysis. To evaluate the dispersion of the distributions we will look at the interquartile range (IQR) which is computed as *IQR* = *Q*3 − *Q*1, being Q1 and Q3 the first and third quartiles of the distribution, respectively. The IQR is a measure of the spread of data that is robust against extreme values. It provides valuable information about the variability of the central portion of a distribution and is helpful for identifying potential outliers. Besides, a comprehensive examination of the outcomes comparing the reflectance measurements from both cameras is presented in Section 5 of the Supplementary Materials.

### 3.3 Identification of Chromophores in Absorbance Measurements

After studying the similarity of the reflectance measurements between the HS cameras in Subsec. 3.1, we now attempt to identify the presence of any of the chromophores mentioned in Subsec. 2.6 within the measured absorbances. Table S3 presents SAM values resulting from the comparison between mean absorbance spectral signatures acquired using both HS cameras and the absorption coefficient spectra of the chromophores discussed in Subsection 3.1. The wavelength of the peaks of interest, the respective analyzed spectral ranges, and the number of bands considered are detailed. Notably, the comparison is made with data obtained from the linescan cameras in the Visible and Near-Infrared (VNIR), using all 369 wavelengths measured by the camera, and NIR regions for the snapshot and linescan cameras as well. For this latter case, we use the 25 overlapping bands between the two cameras, indicated in Table S1. To obtain the SAM values, we extracted the peak wavelength of interest and the fifteen wavelengths on each side of it for the chromophore under analysis. This ensures that the extracted bands correspond to the wavelength of interest and its nearby range. We have chosen to search for a maximum of 15 wavelengths on each side of the peak, as this allows us to extract spectral signatures with sufficient information. We then find the closest corresponding wavelengths between the selected chromophore bands and those measurable by each camera. It is important to note that it will not always be possible to use all 31 bands from the chromophore to identify a peak. This is because the spectral resolution of the cameras is lower than that used to measure the chromophores, hence, there will generally be fewer camera bands than those measured for each chromophore. For example, if the chromophore peak has a very narrow bandwidth, such as that from Cyt. b in its reduced state at *λ* = 555 nm, there will not be many camera bands capable of measuring it. This also applies to searching for the closest snapshot camera wavelengths in the chromophores. In addition, we have included the mean spectral signatures of *A* measured by the cameras together with the *μ*_*a*_ spectral signatures of the chromophore peaks analysed in the Supplementary Materials from Fig. S4 to Fig. S19, allowing the reader to visually check the measurements.

## 4 Discussion

This work compares a HS snapshot camera with a linescan camera in the 659.95 to 951.42 nm range. Although similar studies have employed spectrometer measurements as reference to address comparison between HS cameras,^23^ using such instrument was not feasible in this work since it would have required an sterilization process and place the spectrometer close to an *in-vivo* tissue during multiple surgical procedures. Therefore, the linescan camera was considered to be the reference because it captures 15 times more spectral data and nearly 4 times more spatial data than the snapshot camera. Moreover, a similar linescan camera has already been used as an intraoperative tool for *in-vivo* brain tumor classification with great results in the 400 to 1000 nm range.^15^ Visual analysis were made by comparing the mean spectral signatures reflectance of both cameras in two scenarios, with and without data normalization. Furthermore, objective comparisons were made for both cases by computing four similarity metrics, SAM, GFC, and RMSE. Results have been illustrated in distributions to verify how they spread across the 10 patients used for this study. We computed the IQR for each distribution to study the similarity of both cameras in the two previously described scenarios. Additionally, we attempted to identify several chromophores in the absorbance measured with both cameras. The absorbances were obtained, for each patient, using the gathered reflectance as specified in Subsec. 3.1.

We have observed that the snapshot camera measurements are noisier compared to those from the linescan camera. Regardless of the laser emission at *λ* ≈ 850 nm from the LiDAR of the acquisition system, which must be turned on to visualize a live video during surgery, the spectral signatures appear less smoother than those obtained with the linescan camera. This is evident from the mean reflectance spectral signature of the snapshot, which shows several peaks and valleys along the spectral range from 659.95 to 950.64 nm, which could be due to the noise introduced by the sensor. In fact, the Fabry-Pérot sensor of the snapshot with 25 filters have been characterized in detail by Hahn et. al.,^22^ concluding that the correction matrix provided by the manufacturer is insufficient to reconstruct the spectrum without introducing large measurement errors. Furthermore, the irregularities found in the sensor are present across the whole sensor, hence, in the entire spectral range of the camera. Although Hahn et. al. propose to create an individual matrix after characterizing the camera, a dedicated optical system is required which was not available for this study. To mitigate this issue, previous works by Muhle et. al. using the same snapshot camera model averaged 10 captures for organ transplantation purposes.^23^ However, we could not adopt the same method for imaging *in-vivo* tissue because multiple captures would yield varying spatial measurements due to the brain movement caused by the heartbeat. Despite the noise introduced in the snapshot camera measurements, the trends of the spectral signatures are similar to those obtained with the linescan camera. In general, both cameras seem to agree that the pathological tissue provides less reflectance than healthy tissue when data is unnormalized. Such behaviour can be seen in the two first columns of Fig. S2.

The errors in the snapshot camera measurements are more pronounced in the spectral signatures in Fig. S2 once data is normalized. This was expected since a min-max normalization was applied to each spectral signature individually, causing errors in measurements to increase since data is scaled from the 0 to 1 range. This behaviour is illustrated within the spectral signatures in the two right columns of Fig. S2, where the continuous lines have a noiser trend than the dashed spectral signatures coming from the linescan camera. In consequence, normalized data from both cameras appear to be less similar from each other than when reflectance data is unnormalized. This fact is confirmed after analyzing the distributions with the SAM, GFC and RMSE metrics, which are used to address the comparison of the cameras. Generally, distributions using normalized data have higher IQR values since they are more spreaded than those distributions with unnormalized data. For example, IQR values from unnormalized data for the healthy tissue are 0.023, 0.001, and 0.061 for the SAM, GFC and RMSE metrics, respectively, whereas the results from normalized data are 0.119, 0.006 and 0.083 for the same metrics.

Expected differences in reflectance intensity, as seen in Fig. S2, could also arise due to variations in lighting angle and working distance. These variations are traditionally corrected using a white reference image (*I*_*white*_ from Eq. 1), obtained from a flat calibration board.^15, 18^ However, the 3D structure of the brain and the inherent organ texture variations can cause discrepancies in spectral characteristics due to deviations in illumination and working distance across the surface.^36^ Although unnormalized data make spectral measurements very similar for both cameras, as presented in Fig. 5 (e) and (f), the spectra is very steady throughout the spectral range under analysis. Hence, the presence of absorption peaks, like the one that Hb has at *λ* = 756 nm, might be hardly noticeable with unnormalized data. Normalized data seem to illustrate better the presence of Hb or blood with local minimums at *λ* ≈ 756 nm, as presented in Fig. 6 (e) and (f). This behaviour is found when analyzing the SAM values in Table S3 to identify such absorption peak in the absorbance of the cameras, where normalized data has 0.03 less SAM than that obtained with unnormalized data, regardless of the tissue. However, the SAM values obtained with the snapshot cameras are higher with normalized data than with unnormalized data. This might be due to the noise introduced by the sensor, as already explained previously. Moreover, observations show how the contribution of Hb is slightly higher in pathological tissue than in healthy tissue at *λ* = 756 nm, which may be related to increased perfusion of tumor tissue, especially in high-grade tumors, or may even be related to lack of oxygen to brain tissue or tumor hypoxia due to abnormalities in tumor vessel structure.^37^ Such behaviour was found in other studies using HSI for *in-vivo* human brain^29^ and might also indicate hypoxia from glioma cells.^38^ The analysis conducted in Subsection 3.3 aimed to identify any chromophore absorption peaks in the absorbance measurements of the cameras. The results indicate overoptimistic SAM values for most peaks, which do not seem to correlate with the spectra in Fig. S4 to Fig. S19. However, the absorbance measurements of the linescan camera might indicate the presence of four peaks, as shown by the SAM values in Table S3 and their corresponding spectra in Fig. S5, Fig. S14, Fig. S15, and Fig. 19. These peaks correspond to an absorption peak of oxidised Cyt. b at *λ* = 422 nm, two peaks of HbO2 at *λ* = 542 nm and *λ* = 576 nm, and the water absorption peak at *λ* = 976 nm. Regardless of the tissue under analysis and the data used, the values we obtained are SAM ≈ 0.235 for the peak at *λ* = 422 nm, SAM ≈ 0.210 for the peak at *λ* = 542 nm, SAM ≈ 0.110 for the peak at *λ* = 576 nm, and SAM ≈ 0.235 for the peak at *λ* = 976 nm. This statement makes sense because the first three peaks have the highest absorption coefficient values, which means they could potentially be measured. Although these peaks are the most absorbent and might help during tumor detection, further research is needed to evaluate if the snapshot camera employed can detect pathological tissue in the 659.95 to 950.64 nm range.

## 5 Conclusions

In this study, we have compared and analyzed two different HS cameras because of their potential to be used for intraoperative brain tissue identification. Specifically, we have used images from 10 *in-vivo* human patients with different pathologies. Measurements show how the snapshot camera with less spectral and spatial resolution can capture a similar spectral behaviour than that obtained with the linescan camera. Although the linescan camera has almost 3 times more spectral resolution than the snapshot, it required 1 minute and 40 seconds to scan a single *in-vivo* human brain image. Hence, it is not suitable for real-time solutions compared with the snapshot camera, which has already been used in real-time solutions for brain tumor classification.^19^ Moreover, objective comparisons were made in the shared spectral range of both cameras between 659.95 to 951.42 nm using four similarity metrics: SAM, GFC, RMSE, and PCC. Results with unnormalized data show high similarity between the reflectances captured with the cameras in the aforementioned spectral range for either healthy or pathological tissues. However, due to the noise introduced by the snapshot mosaic sensor,^22^ the similarity between cameras are reduced once data is normalized. For example, the SAM metric shows that there is reduced dispersion and high similarity between cameras for pathological samples, with an IQR value of 9.68% for normalized data, compared to an IQR value of 2.38% for unnormalized data. This behaviour is consistent also for both GFC and RMSE, irrespective of the type of tissue under inspection. Differences in similarity between cameras may be attributed to errors that arise from the independent normalization applied to each spectral signature to minimum and maximum values between 0 and 1. Even though noiseless measurements from the snapshot camera could be obtained by averaging multiple images,^22, 23^ such procedure is not feasible during *in-vivo* brain surgeries due to the heart beating, which pumps blood to the brain causing it to move. Furthermore, we studied the ability of both cameras to identify several tissue chromophores in their measurements. In particular, we attempted to identify Hb, HbO2, fat, water, and several cytochromes by converting the measured reflectance of the cameras to absorbance, as specificed in Subsec. 3.1. Such task is done by trying to identify relevant peaks of the previous chromophores. For that, we apply the SAM metric as it is the only one that only considers the shape of the spectra. Furthermore, the identification of chromophores was also conducted through a subjective inspection of the absorbance spectra measured with the cameras in comparison to the absorption coefficients of the chromophores. Out of the twenty one peaks analysed, only five could potentially be identified by the snapshot camera as most of them are present in the visible spectrum, specifically from the 400 to 625 nm spectra. However, the snapshot camera encountered difficulties in identifying any of those five peaks, which are from oxidised Cyt. c at *λ* = 695 nm, Hb at *λ* = 756 nm, and three fat peaks at *λ* = 756 nm, *λ* = 830 nm, and *λ* = 930 nm. Such difficulties could be due to their low absorption coefficient values compared to those in the visible spectrum and the low spectral resolution of the camera. Nonetheless, we observed that the linescan camera detected four absorption peaks, which corresponded to three different chromophores present in its absorbance measurements. These peaks correspond to the oxidised Cyt. b peak at *λ* = 422 nm, to two peaks of HbO2 at *λ* = 542 nm and *λ* = 576 nm, and to a water peak at *λ* = 976 nm. Regardless of the tissue and data used, the obtained SAM values between the absorbance of the camera and the absorption coefficient of the chromophores were approximately 0.235, 0.210, 0.110, and 0.100, respectively. These values suggest high similarities between the spectra and the possible presence of the mentioned chromophores in the absorbance measurements.

All things considered, the snapshot camera can provide reasonable measurements to describe brain tissue behaviour when compared with the typical linescan cameras used for brain tumor detection.^15, 29, 39, 40^ Likewise, the snapshot camera offers great opportunities to provide real-time solutions as employed in other studies.^19^ However, combining multiple snapshot cameras to increase the spectral range can lead to a better reconstruction of the spectral behaviour of biological tissues as shown in other works.^23^ Therefore, this study shows the potential use of snapshot cameras for *in-vivo* brain tissue identification. Moreover, similar spectral measurements from both cameras were obtained, suggesting the combination of data from both cameras to train classification models and enhance *in-vivo* brain tumor classification.

## Data Availability

All the in-vivo hyperspectral human brain data used in this study are from the Slim Brain database. Note that access must be granted, under reasonable request, before downloading the data.

https://slimbrain.citsem.upm.es/

## 7 Acknowledgments

The authors would like to thank neurosurgeons and staff of the Hospital Universitario 12 de Octubre as well as the contributors and members of the TALENT-HIPSTER (High Performance Systems and Technologies for E-health and fish Farming) (PID2020-116417RB-C41) research projects, funded by the Spanish Ministry of Science and Innovation.

## 8 Code, Data, and Materials Availability

All the *in-vivo* hyperspectral human brain data used in this study are from the Slim Brain database, which is available at https://slimbrain.citsem.upm.es/. Note that access must be granted, under reasonable request, before downloading the data.

**Alberto Martín-Pérez** acquired a Master’s Degree in Internet of Things (IoT) and a Bachelor’s Degree in Sound and Image Engineering from Technical University of Madrid (UPM), achieving these qualifications in 2021 and 2020, respectively. Presently, he is pursuing a PhD at UPM with the Electronic and Microelectronic Design Group (GDEM) in the Software Technologies and Multimedia Systems for Sustainability (CITSEM) Research Center. His research pursuits center around the utilization of Machine Learning algorithms for the classification of in-vivo human brain tumours through hyperspectral imaging. Furthermore, he aims to enhance classification methodologies through the application of spatial frequency domain imaging for his doctoral studies.

**Alejandro Martínez de Ternero** received his master degree in Internet of Things (IoT) and his bachelor degree in Telematics from Universidad Politécnica de Madrid (UPM), Spain, in 2022 and 2021 respectively. He is currently a PhD student at the Electronic and Microelectronic Design Group (GDEM) in the Software Technologies and Multimedia Systems for Sustainability (CITSEM) Research Center, UPM. His research is focused on modeling the light-tissue interactions through Monte Carlo and Radiative Transfer modeling for optical property estimation. Using the information captured by hyperspectral cameras and Spectral Unmixing techniques to expedite classification of tissue types.

**Prof. Alfonso Lagares** (male) is Professor in Neurosurgery in Universidad Complutense de Madrid and Head of Department of Neurosurgery in Hospital 12 de Octubre. He received his PhD in Neuroscience in 2004 in Universdad Autonoma de Madrid achieving Doctorate Extraordinary Prize. He is the coordinator of a research group in the research Institute imas12. His lines of investigation includes: 1) Prognostic models in traumatic brain injury and subarachnoid hemorrhage; 2) New tools for in different neurosurgical pathologies including radiological tools such as volumetric CT, conventional MR as well as diffusion and MR for assessing white matter integrity in diffuse axonal injury; 3) Discovery of new biomarkers in the diagnosis and prognosis of head injury; 4) Design of new tools for improving tumour resection. He is author or co-author of more than 250 papers, received more than 4700 citations, and has participated in 20 competitive projects.

**Dr. Eduardo Juárez** (male) received his PhD degree from the É cole Polythecnique Fédéral de Lausanne (EPFL) in 2003. From 1994 to 1997, he worked as researcher at the Digital Architecture Group of UPM and was a visiting researcher at the ENST, Brest (France) and the University of Pennsylvania, Philadelphia (USA). From 1998 to 2000, he worked as Assistant at the Integrated Systems Laboratory (LSI) of the EPFL. From 2000 to 2003, he worked as Senior Systems Engineer at the Design Centre of Transwitch Corp. in Switzerland, while continuing his research towards the PhD at the EPFL. In December 2004, he joined the Universidad Politecnica de Madrid (UPM) as a postdoc. Since 2007, he is Associate Professor at UPM. His research activity is mainly focused on (1) hyperspectral imaging for health applications, (2) real-time depth estimation and refinement and (3) heterogeneous high performance computing. He is co-author of one book and author or co-author of more than 100 papers and contributions to technical conferences. He has participated in more than 15 competitive research projects and 20 non-competitive industrial projects.

**César Sanz** received the PhD degree in Telecommunication Engineering in 1998, from the Universidad Politécnica de Madrid (UPM). Since 1985, he has been a faculty member at the UPM, where he is currently a Full Professor. He has been the Director of the ETSIS de Telecomunicación, UPM, from 2008 to 2017. He leads the Electronic and Microelectronic Design Group (GDEM) since 1996, involved in R&D projects with private companies and public institutions. He is author and/or co-author of 10 books, an international patent and more than 100 papers and contributions to technical conferences. He has participated in more than 80 R&D projects. Since 2021, he is the director of CITSEM research center at UPM. His research interests include electronic design applied to video coding and hyperspectral imaging.

