## Supplementary Materials for "Spectral Analysis Comparison of Pushbroom and Snapshot Hyperspectral Cameras for *In-Vivo* Brain Tissues and Chromophores Identification"

#### 1 Wavelengths shared between hyperspectral cameras

The wavelengths shared between both HS cameras are presented in Table S1. Note that we have used the linescan camera bands that are closest to those measured by the snapshot.

**Table S1** Wavelengths in nm used to compare the snapshot and linescan HS cameras. It includes the absolute difference for each wavelength between cameras, including the mean and standard deviation in nm of all wavelength differences.

| **Snapshot 5×5-mosaic [nm]** | **Pushbroom linescan [nm]** | **\|Difference\| [nm]** |
|---|---|---|
| 659.95 | 660.61 | 0.66 |
| 670.69 | 671.98 | 1.29 |
| 688.90 | 689.85 | 0.95 |
| 702.51 | 702.85 | 0.34 |
| 713.62 | 714.22 | 0.60 |
| 729.65 | 730.47 | 0.82 |
| 740.67 | 741.84 | 1.17 |
| 753.57 | 754.84 | 1.27 |
| 768.10 | 769.46 | 1.36 |
| 780.73 | 780.83 | 0.10 |
| 792.00 | 792.21 | 0.21 |
| 807.38 | 808.45 | 1.07 |
| 817.51 | 818.20 | 0.69 |
| 830.18 | 831.20 | 1.02 |
| 845.03 | 845.82 | 0.79 |
| 855.45 | 855.57 | 0.12 |
| 866.50 | 866.94 | 0.44 |
| 880.23 | 881.56 | 1.33 |
| 891.89 | 892.93 | 1.04 |
| 899.84 | 901.06 | 1.22 |
| 914.48 | 915.68 | 1.20 |
| 922.02 | 922.18 | 0.16 |
| 931.74 | 931.93 | 0.19 |
| 940.95 | 941.67 | 0.72 |
| 950.64 | 951.42 | 0.78 |
| | **Mean ± standard deviation** | 0.78±0.41 |

### 2 Brain images used from both cameras

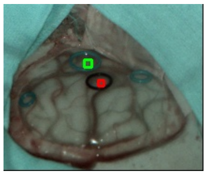
(a) Patient ID177 - Snapshot

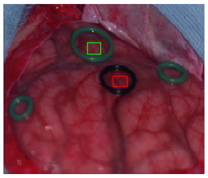
(b) Patient ID177 - Linescan

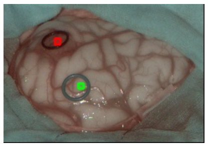
(c) Patient ID183 - Snapshot

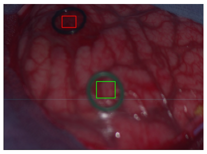
(d) Patient ID183 - Linescan

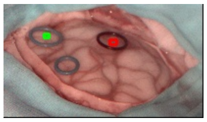
(e) Patient ID184 - Snapshot

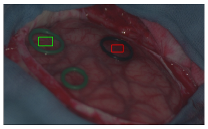
(f) Patient ID184 - Linescan

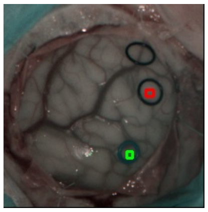
(g) Patient ID185 - Snapshot

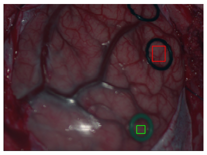
(h) Patient ID185 - Linescan

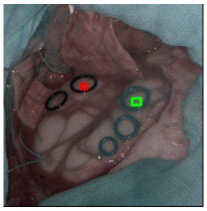
(i) Patient ID190 - Snapshot

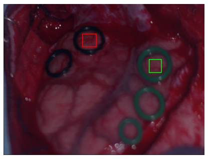
(j) Patient ID190 - Linescan

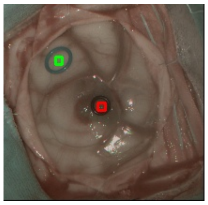
(k) Patient ID192 - Snapshot

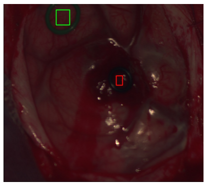
(l) Patient ID192 - Linescan

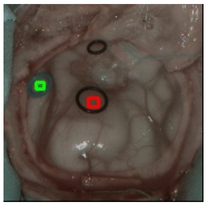
(m) Patient ID193 - Snapshot

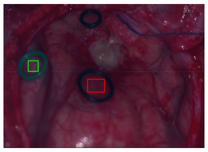
(n) Patient ID193 - Linescan

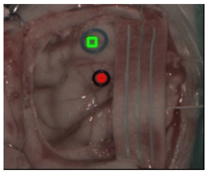
(o) Patient ID194 - Snapshot

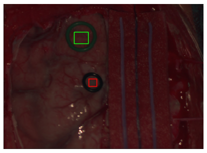
(p) Patient ID194 - Linescan

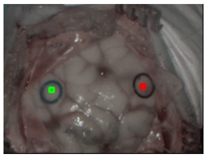
(q) Patient ID201 - Snapshot

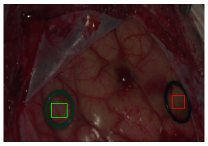
(r) Patient ID201 - Linescan

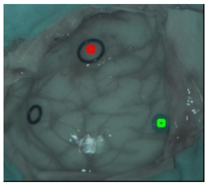
(s) Patient ID203 - Snapshot

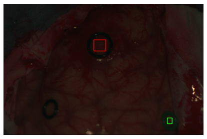
(t) Patient ID203 - Linescan

**Fig S1** Patients used for this study with their identifier. First and third columns are the pseudo RGB with healthy and pathological region of interests (ROIs) for the HS snapshot captures. Namely, the second and forth columns represent the same but for the captures taken with the HS linescan camera.

### 3 Reflectance measurements for each patient

The reflectance measurements obtained with both HS cameras from the 10 *in-vivo* human brains are shown in Fig. S2. The illustrated spectral signatures have been obtained, for every camera measurement, using 25 pixels located inside the rubber rings in the brain captures.

When looking at the unnormalized data, similar spectral signatures are measured with both HS cameras for 8 out of the 10 *in-vivo* human brains. Note how the spectral signatures of both cameras for patients 192 and 193 are less similar compared to other patients for either the healthy or pathological tissues. Furthermore, the healthy tissue measurements from the different patients have values between 0.2 to 0.6 in most cases, excluding patient 192 which has values between 0.4 to almost 1.0. Moreover, the reflectance values for pathological tissues range from 0.1 to 0.4 for most patients, excluding patient 185 whose values are between 0.5 to 0.8. The variations in reflectance or spectral signatures could result from the biological differences of the patients and their distinct brain tumor pathologies.

Overall, the mean spectral signatures of both cameras differ more from each other once the normalization is applied. For instance, the healthy tissue measurements of patient 183 show a 0.3 reflectance difference between the spectral signatures of both cameras, whereas the unnormalized data for the same patient has a variation less than 0.05. Such behaviour can be seen for most patients and tissues except the healthy spectral signatures of patient 192, since it already exhibits great difference in the unnormalized measurements. Additionally, we can see an increase in the standard deviation shown in shaded colors around the mean curves due to the normalization. It is worth noting that the location of each pixel on the curved brain surface implies light variations, leading to slightly different measured reflectances. This, in turn, increases the difference between pixels within the same ROI when normalizing the reflectance to a range between 0 to 1. While measurements between cameras display a consistent trend across patients and tissues, snapshot measurements show more noise after normalization compared to the smoother linescan camera measurements. This results in more pronounced peaks and valleys in the spectrum of the snapshot measurements. However, there is a common peak across most measurements at $\lambda \approx 850$ nm, which is influenced by the infrared of the depth camera used while capturing only with the snapshot camera. Such peak can be seen in patient 203 measurements in Fig. S1. Variations of amplitude of such peak are due to the difference angles at which the patients were captured. By comparing measurements with and without normalization, it is worth noting that the aforementioned peak at $\lambda \approx 850$ nm is more pronounced once the normalization is applied.

The spectral similarity metrics used to compare both cameras patient by patient, including both tissues under analysis, are presented in Table S2.

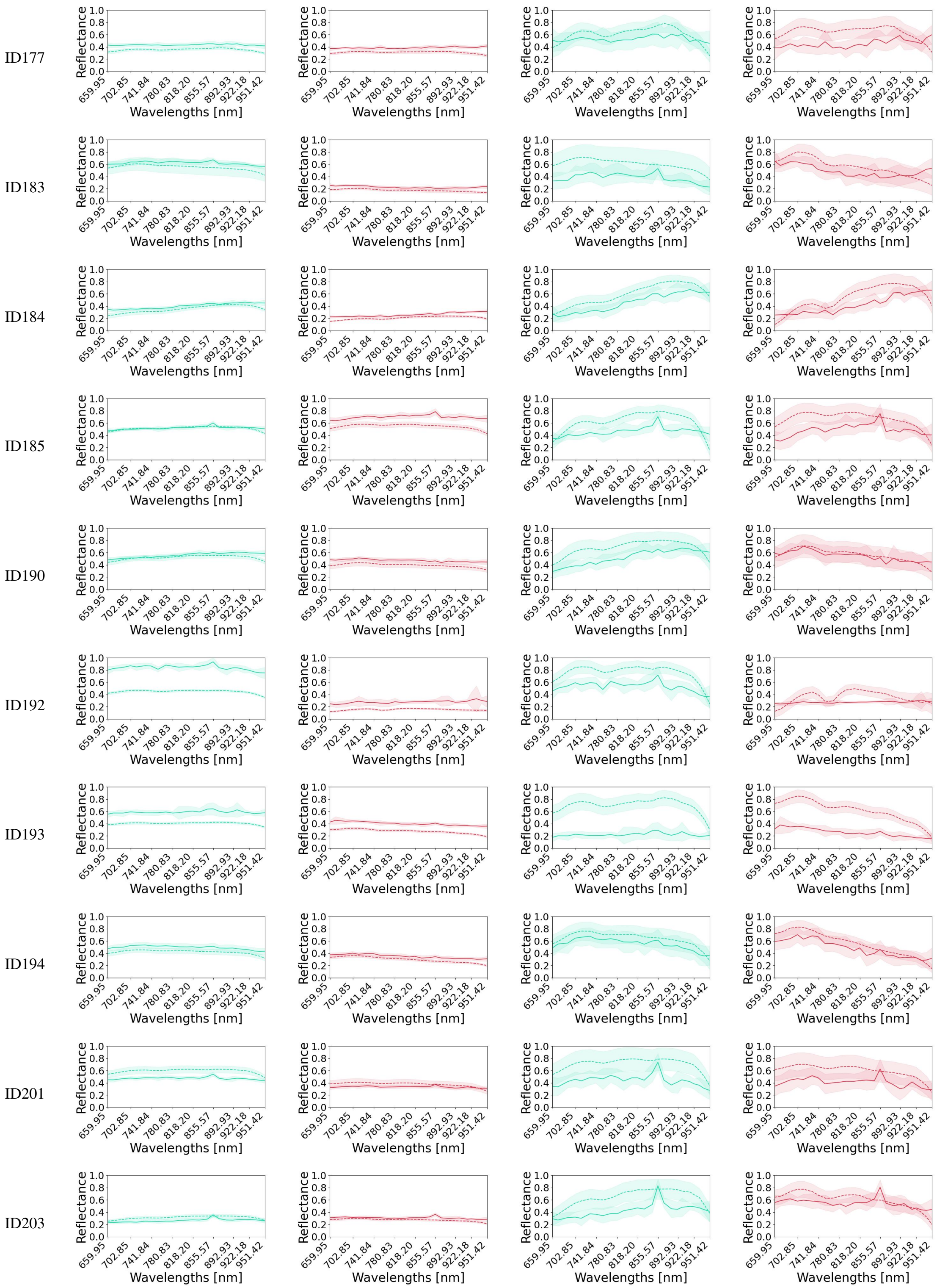


**Fig S2** Mean spectral signatures with standard deviation for every patient in the study. Data comes from the 25 pixels included inside the rubber rings captured with both HS cameras. Continuous curves are from the snapshot camera, while dashed line curves are from the linescan camera. Plots are in green and red to represent the healthy and pathological tissues, respectively. The two columns on the left are the spectral signatures when data has been calibrated and denoised, while in the two columns to the right data has additionally been normalized using a min-max normalization.

**Table S2** Spectral similarity metrics between both HS cameras for every patient. The data used comes from the calibrated and denoised data as well as from the normalized 25 pixels located inside the rubber rings of the captures.

| | | **Calibrated and denoised data** | | | | **Normalized data** | | | |
|---|---|---|---|---|---|---|---|---|---|
| **Patient ID** | **Tissue** | **SAM** | **GFC** | **RMSE** | **PCC** | **SAM** | **GFC** | **RMSE** | **PCC** |
| 177 | Healthy | 0.048 | 0.998 | 0.081 | 0.726 | 0.159 | 0.987 | 0.120 | 0.726 |
| | Pathological | 0.074 | 0.997 | 0.082 | -0.360 | 0.252 | 0.968 | 0.226 | -0.360 |
| 183 | Healthy | 0.055 | 0.998 | 0.077 | 0.788 | 0.122 | 0.992 | 0.224 | 0.788 |
| | Pathological | 0.092 | 0.995 | 0.057 | 0.584 | 0.219 | 0.976 | 0.134 | 0.584 |
| 184 | Healthy | 0.071 | 0.997 | 0.055 | 0.915 | 0.122 | 0.992 | 0.144 | 0.915 |
| | Pathological | 0.092 | 0.995 | 0.060 | 0.704 | 0.253 | 0.967 | 0.174 | 0.704 |
| 185 | Healthy | 0.044 | 0.998 | 0.024 | 0.648 | 0.200 | 0.979 | 0.199 | 0.648 |
| | Pathological | 0.062 | 0.998 | 0.158 | 0.484 | 0.196 | 0.980 | 0.205 | 0.484 |
| 190 | Healthy | 0.055 | 0.998 | 0.051 | 0.649 | 0.168 | 0.985 | 0.176 | 0.649 |
| | Pathological | 0.043 | 0.999 | 0.081 | 0.865 | 0.098 | 0.995 | 0.055 | 0.865 |
| 192 | Healthy | 0.038 | 0.999 | 0.390 | 0.796 | 0.115 | 0.993 | 0.235 | 0.796 |
| | Pathological | 0.099 | 0.995 | 0.128 | 0.270 | 0.256 | 0.967 | 0.118 | 0.270 |
| 193 | Healthy | 0.039 | 0.999 | 0.185 | 0.525 | 0.144 | 0.989 | 0.485 | 0.525 |
| | Pathological | 0.071 | 0.997 | 0.127 | 0.916 | 0.144 | 0.993 | 0.362 | 0.916 |
| 194 | Healthy | 0.029 | 0.999 | 0.074 | 0.933 | 0.063 | 0.997 | 0.093 | 0.933 |
| | Pathological | 0.075 | 0.997 | 0.057 | 0.954 | 0.113 | 0.993 | 0.102 | 0.954 |
| 201 | Healthy | 0.042 | 0.999 | 0.123 | 0.680 | 0.147 | 0.989 | 0.276 | 0.680 |
| | Pathological | 0.076 | 0.997 | 0.048 | 0.630 | 0.156 | 0.987 | 0.198 | 0.630 |
| 203 | Healthy | 0.074 | 0.997 | 0.051 | 0.645 | 0.196 | 0.980 | 0.235 | 0.645 |
| | Pathological | 0.062 | 0.998 | 0.039 | 0.592 | 0.183 | 0.983 | 0.119 | 0.592 |

### 4 Analysis of the absorbance measurements to identify chromophores' peaks

To identify chromophores' peaks, we will first look for their maximum absorption peaks. Such peaks are indicated with circular markers in Fig. S3. The absorption coefficients of those peaks are given in the left list below the plot for their corresponding wavelengths. In addition, we will also try to look for the presence of other absorption peaks that we have found to be of interest. These peaks are indicated in Fig. S3 with diamond markers and are given in the right list below the plot. As can be seen in the figure, most of the absorption peaks could be identified within the visible (VIS) range measured by the linescan camera. However, certain peaks in the near infrared (NIR) spectrum could be identified with the snapshot camera.

Analyzing the absorbance measured with the linescan camera in Fig. 5 (a) and Fig. 5 (b), there is an absorbance peak at $\lambda \approx 425$ nm in both tissues. Such peak is located inside the black dashed rectangle (A) and might be influenced by both absorption peaks of HbO2 and Hb at $\lambda = 414$ nm and $\lambda = 432$ nm, respectively. However, it also might be influenced by the absorption peak of Cyt b in its reduced state at $\lambda = 422$ nm, as seen in Fig. 5 (c) and Fig. 5 (d). Likewise, the two absorption peaks of HbO2 at $\lambda = 542$ nm and $\lambda = 572$ nm, located inside the black dashed rectangles (B), seem to be detected by the linescan camera for either healthy or

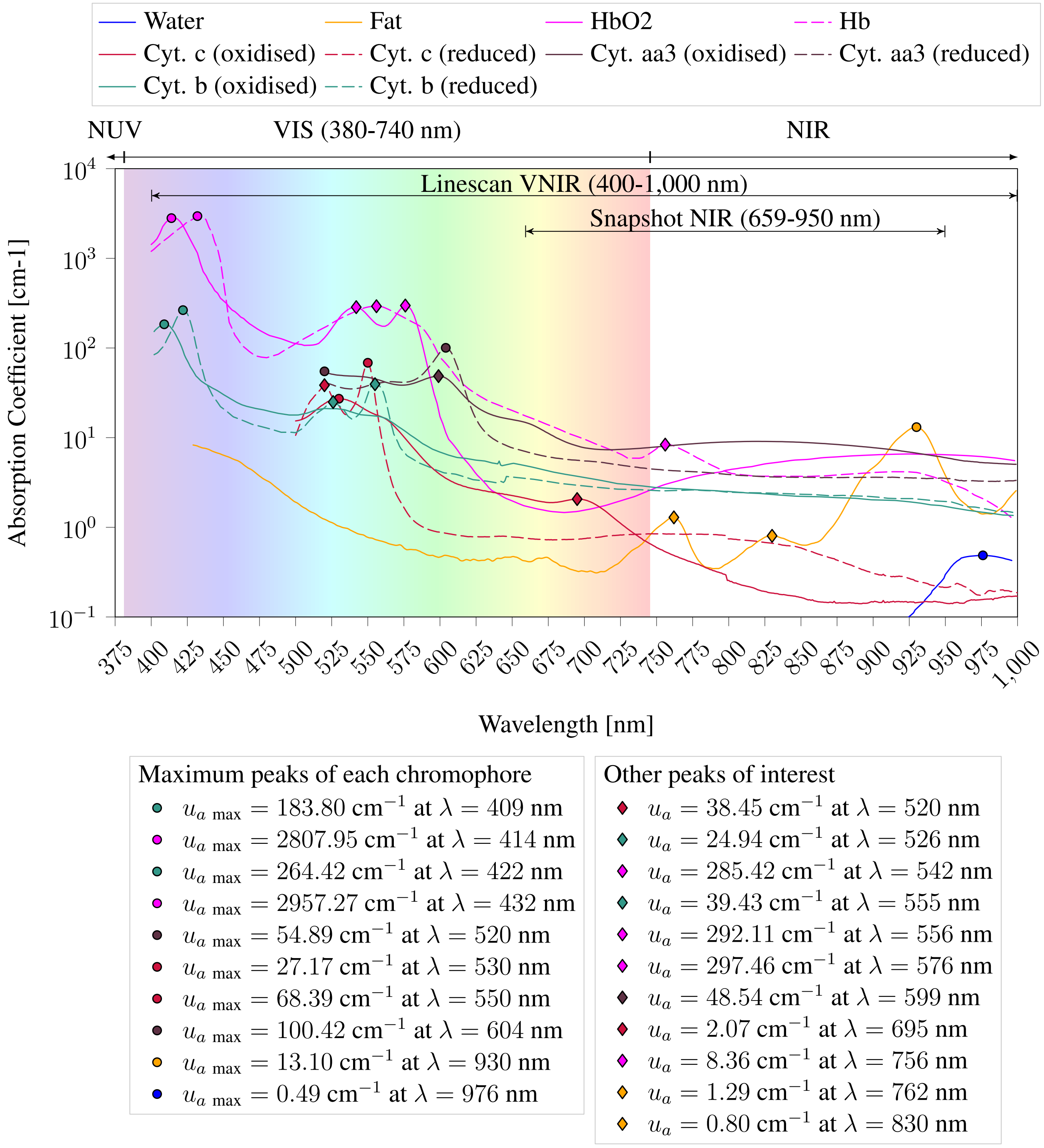


**Fig S3** Absorption coefficient spectra for each tissue chromophore under analysis. For each chromophore the maximum value of the absorption coefficient at the corresponding wavelength is given. Additionally, other absorption coefficient peaks of interest are also listed. The visible (VIS) spectrum is shown with a rainbow shadow in the 380 to 740 nm range, while the near ultraviolet (NUV) and near infrared (NIR) spectrums are shown on the sides. In addition, the spectra that each HS camera can measure are specified within the plot.

pathological tissues. This behaviour was expected, since similar studies have observed the same peaks in their measurements when using another HS linescan with similar specifications to capture *in-vivo* brain HS images.[1,2] Moreover, of all the cytochromes absorption peaks in rectangles (B), the only one that could influence the spectral signatures of the linescan camera is the peak of the Cyt c in its reduced form at $\lambda = 550$ nm. This could be true since such cytochrome is expected to be found in high concentrations in glioma tumour cells,[3] which may have been captured in the brains of some of the patients in this study. The previous observations can also be seen when data is normalized in all the plots of Fig. 6. Furthermore, pathological tissues seem to absorb more than the healthy tissues in the entire 400 to 1,000 nm range, a fact already observed in other studies.[4–6] This pattern is present when data have not been normalized in Figs. 5 (a)-(f). Specifically, the pathological tissues absorbs 20.27% more than the healthy tissues. Such value is computed by subtracting the green and red dashed lines curves from Fig. 5 (a) and Fig. 5 (b), band by band, and compute the mean of all bands. If we instead compute the same difference but for the snapshot camera measurements in the spectrum range from 659.95 to 950.64 nm, the pathological tissue absorbs 18.97% more than healthy tissue. This is again computed by subtracting the green and red continuous lines curves from Fig. 5 (e) and Fig. 5 (f), band by band, to then compute the mean. Following the same procedure for the linescan and snapshot measures when data is normalized as in Figs. 6 (a) to (f), we see again that the pathological tissue absorbs more light than healthy tissue in the snapshot spectral range. Concretely, 11.02% and 6.87% for the snapshot and linescan measurements, respectively. Furthermore, analyzing the same difference for the entire spectrum measured by the linescan, as presented in Fig. 6 (a) and Fig. 6 (b), we obtain that the pathological tissue absorbs 11.84% more than the healthy tissue. Thus, these two latest analysis with the linescan measures show a 4.97% increase in absorption when the visible range is included. Such increase probably has to do with the presence of the peaks with highest absorption for Hb and HbO2 in the 400 to 600 nm range, or more specifically, those peaks inside the (A) and (B) rectangles. In addition, both snapshot and linescan measurements for healthy and pathological tissues appear to be influenced by the Hb absorption peak at $\lambda = 756$ nm. This is indicated in plots (a), (b), (e), and (f) from either Fig. 5 or Fig. 6 with the black dashed rectangle (C), being most noticeable when data is normalized as in Fig. 6 (e) and Fig. 6 (f).

Finally, only the linescan measurements seem to capture the water absorption peak at $\lambda = 976$ nm, as presented with the black dashed rectangle (D) in Fig. 5 (a)-(b) and Fig. 6 (a)-(b). The snapshot camera can not measure such peak since it is only able to measure up to 950.64 nm. Nevertheless, the absorption peak of fat at $\lambda = 930$ nm might not be captured by any of the cameras since there is no noticeable peak found in the spectral measurements at that wavelength. The reason behind could be a combination of the low light intensity provided by the halogen lamp after 900 nm, the low sensitivity of the cameras at highest wavelengths, and the low absorption peak value of fat at $\lambda = 930$ nm. The value of such peak is 13.10 $cm^{-1}$, which is almost 225-fold smaller compared to the highest peaks of HbO2 and Hb with values of 2807.94 $cm^{-1}$ and 2957.26 $cm^{-1}$, respectively. Moreover, fat may be located in deeper layers of the tissue where HSI is unable to interrogate.

### 5 Analysis of the reflectance measurements to compare both cameras

Apart from the SAM, GFC, and RMSE metrics, we also analyzed the Pearson Correlation Coefficient (PCC) between spectral signatures since it was the metric used in a similar study which assessed the comparison of spectral cameras for image-guided organ transplantation.[7] However, in this study it did not describe properly the differences in the spectral signatures between unnormalized and normalized data. Specifically, the results were almost identically for both cases even though we double checked the computation process of PCC and found no programming errors. Thus, PCC seems to be overoptimistic even when SAM, GGC, and RMSE clearly identified variations between the cameras when data was normalized.

Analyzing the SAM metric in Fig. 7 (a) and Fig. 7 (b), the distributions for healthy and pathological tissues are more concentrated when data are unnormalized than when normalized. For instance, the healthy tissue measurements in green present an IQR value of 0.027 in Fig. 7 (a), whereas in (b) the distribution has an IQR value of 0.067. Distributions with the pathological measurements in red show IQR values of 0.023 and 0.119 in Fig. 7 (a) and Fig. 7 (b), respectively. Observing the GFC results, the distributions for healthy and pathological tissues are less dispersed when data are not normalized than when data are normalized, as seen in Fig. 7 (c) and Fig. 7 (d), respectively. The IQR values for the healthy tissue distributions are 0.001 for Fig. 7 (c) and 0.006 for Fig. 7 (d), whereas for the pathological tissue distributions, the IQR values are 0.002 and 0.022 for the same figures, respectively. When we examine the RMSE metric results, we noticed again a consistent pattern similar to that observed with the SAM and GFC results, regardless of the tissue. In this pattern, the distributions tend to be more tightly clustered when the data remains unnormalized. On one hand, in Fig. 7 (e) and Fig. 7 (f), we observe that the IQR values are 0.061 and 0.083 for the healthy tissue in both unnormalized and normalized distributions, respectively. On the other hand, the IQR values are 0.058 and 0.085 for the pathological tissue distributions in the same figures for the unnormalized and normalized data distributions, respectively. Note that the two outliers indicated with the black diamond marks in Fig. 7 (e) and Fig. 7 (f) correspond to the patients 192 and 193, respectively. These patients are already identified as possible outliers in Fig. S2 since the spectral signatures from both cameras were less similar than those compared to other patients. Examining the unnormalized spectral signatures for the healthy tissue in Fig. S2 for patient 192, it is evident why the RMSE obtained for such patient is higher than the rest of the RMSE values obtained for the other patients, thereby causing it to be an outlier. This can be double checked by looking at the RMSE results obtained for every patient in Table S2. Additionally, the same behaviour happens for patient 193 for the normalized healthy and pathological tissues distributions in Fig. 7 (f), whose outliers correspond to patient 193.

Nonetheless, the pattern related to the increase of dispersion in the distributions when normalizing the data is not seen when observing the obtained results for the PCC metric presented in Fig. 7 (g) and Fig. 7 (h). In this case, the distributions seem to be unaffected by the normalization since the results are almost the same (we could only see differences in the 16th decimal). The IQR value for healthy tissue distributions are 0.140 and 0.146 for the unnormalized and normalized distributions, respectively, whereas for the pathological tissue distributions the IQR values are 0.313 and 0.316, also for the unnormalized and normalized distributions in Fig. 7 (g) and Fig. 7 (h), respectively.

### 6 Spectral comparison of relevant chromophore absorption coefficient peaks with absorbance spectral signatures of the hyperspectral cameras

The comparison is numerically done employing the SAM metric to compare the shapes of the spectral signatures, presented in Table S3. Moreover, Fig. S4 to Fig. S19 serve as visual guide to potentially identify absorption coefficient peaks in the measurements taken with the hyperspectral cameras. The black circle indicates the maximum peak of the chromophore, while black diamond indicates an interesting absorption peak.

For instance, considering the Cyt. aa3 (reduced) peak wavelength at $\lambda = 604$ nm, the SAM values for healthy and pathological tissues, both unnormalized and normalized, are presented as 0.3019 and 0.3571, and 0.3021 and 0.3877, respectively. However, such peak does not seem to be visible in the absorbance of the linescan camera after inspecting Fig. S4 (a) and (b). Similarly, for Cyt. aa3 (oxidized) at $\lambda = 520$ nm and $\lambda = 599$ nm peaks, the corresponding SAM values suggest the possible presence of the chromophore. However, Fig. S5 (a), (b), (c), and (d) do not show any evidence of the presence of such peaks. Furthermore, Cyt. b in its reduced form is analyzed at peaks of $\lambda = 422$ nm, $\lambda = 526$ nm, and $\lambda = 555$ nm. Contrastingly, oxidized Cyt. b at $\lambda = 409$ nm is evaluated with its SAM values under the given spectral conditions. Although the SAM values suggest the presence of oxidized Cyt. b in the measurements, a visual inspection of Fig. S6, Fig. S7, and Fig. S8 might indicate that only the peak at $\lambda = 422$ nm is present in the absorbance. Moving to the reduced Cyt. c, the analyzed peaks at $\lambda = 520$ nm and $\lambda = 550$ nm show SAM values below 0.26, regardless of the data and the tissue, potentially indicating the presence of the chromophore. However, a visual inspection of Fig. S9 reveals that the absorbance spectra and the absorption coefficient spectrum of the chromophore follow different trends. Therefore, it is unlikely that the peak of that chromophore was present in the measurements. Analyzing the oxidised Cyt. c SAM values with the spectra compared in Fig. S10 at $\lambda = 530$ nm and $\lambda = 695$ nm, it does not seem that both peaks of the chromophore could be present in the measurements taken by any of the cameras and wavelengths used. Additionally, the Hb peaks at $\lambda = 432$ nm, $\lambda = 556$ nm, and $\lambda = 756$ nm are examined with their corresponding SAM values and spectra using Fig. S11 and Fig. S12. Although we obtained low SAM values for the $\lambda = 556$ nm, and $\lambda = 756$ nm peaks, the visual inspection might indicate that none of the peaks are present in the measurements of the cameras. However, after observing the HbO2 peaks at $\lambda = 542$ nm, $\lambda = 576$ nm, we can see a correlation between the SAM values below 0.24 obtained with the possible presence of those peaks in the absorbances presented in Fig. S14 and Fig. S15, regardless of the tissue under inspection. Analyzing the fat, we see that the SAM values obtained for the three peaks at $\lambda = 762$ nm, $\lambda = 830$ nm, and $\lambda = 930$ nm seem to indicate their presence in the absorbance of the cameras since most values are below 0.28. Nonetheless, it seems difficult to identify these peaks in the spectra of the cameras presented in Fig. S16, Fig. S17, and Fig. S18. Finally, it is worth noting the possible correlation between the low SAM values below 0.13 for the water peak at $\lambda = 976$ nm, given that the spectra shown in Fig. S19 might indicate the presence of such absorption peak in the absorbance measured by the linescan camera.

**Table S3** SAM values obtained after comparing the mean absorbance spectral signatures measured with both cameras with the absorption coefficient spectra of the chromophores under inspection.

| | | | | | Unnormalized data | | Normalized data | |
|---|---|---|---|---|---|---|---|---|
| **Chromophore** | **Peak [nm]** | **Analyzed range $\lambda$ [nm]** | **Number of bands** | **Camera** | **Healthy** | **Pathological** | **Healthy** | **Pathological** |
| **Cyt. aa3 (reduced)** | 604 | 589.13 - 618.37 | 19 | Linescan (VNIR) | 0.3019 | 0.3021 | 0.3571 | 0.3877 |
| **Cyt. aa3 (oxidised)** | 520 | 519.27 - 535.51 | 11 | Linescan (VNIR) | 0.0668 | 0.0594 | 0.0861 | 0.0821 |
| | 599 | 584.25 - 613.49 | 19 | | 0.1509 | 0.1449 | 0.2470 | 0.2791 |
| **Cyt. b (reduced)** | 422 | 511.14 - 540.39 | 19 | Linescan (VNIR) | 0.3887 | 0.3892 | 0.3745 | 0.3688 |
| | 526 | 540.39 - 569.63 | 19 | | 0.1602 | 0.1540 | 0.1755 | 0.1699 |
| | 555 | 402.29 - 423.41 | 14 | | 0.3733 | 0.3768 | 0.3741 | 0.3823 |
| **Cyt. b (oxidised)** | 409 | 407.17 - 436.41 | 19 | Linescan (VNIR) | 0.2339 | 0.2311 | 0.2495 | 0.2513 |
| **Cyt. c (reduced)** | 520 | 504.64 - 535.51 | 20 | Linescan (VNIR) | 0.2609 | 0.2561 | 0.2756 | 0.2728 |
| | 550 | 535.51 - 564.76 | 19 | | 0.5671 | 0.5705 | 0.5643 | 0.5703 |
| **Cyt. c (oxidised)** | 530 | 514.39 - 545.26 | 20 | Linescan (VNIR) | 0.0800 | 0.0741 | 0.0945 | 0.0861 |
| | 695 | 680.11 - 709.35 | 19 | Linescan (VNIR) | 0.0487 | 0.0464 | 0.0975 | 0.0921 |
| | | 671.98 - 714.22 | 4 | Linescan (NIR) | 0.0639 | 0.0604 | 0.2220 | 0.1929 |
| | | 688.90 - 713.62 | 3 | Snapshot (NIR) | 0.0394 | 0.0307 | 0.0662 | 0.0405 |
| **Hb** | 432 | 402.29 - 462.40 | 31 | Linescan (VNIR) | 0.4367 | 0.4588 | 0.3917 | 0.4137 |
| | 556 | 525.76 - 585.88 | 31 | | 0.1382 | 0.1490 | 0.1350 | 0.1572 |
| | 756 | 725.60 - 785.71 | 31 | Linescan (VNIR) | 0.1325 | 0.1315 | 0.1013 | 0.1085 |
| | | 730.47 - 780.83 | 5 | Linescan (NIR) | 0.1233 | 0.1141 | 0.1444 | 0.1253 |
| | | 729.65 - 780.73 | 5 | Snapshot (NIR) | 0.1411 | 0.1220 | 0.1573 | 0.1033 |
| **HbO2** | 414 | 400.67 - 444.53 | 23 | Linescan (VNIR) | 0.4076 | 0.4089 | 0.3840 | 0.3760 |
| | 542 | 512.77 - 571.25 | 31 | | 0.2266 | 0.2352 | 0.2076 | 0.2111 |
| | 576 | 545.26 - 605.37 | 31 | | 0.4097 | 0.4251 | 0.3493 | 0.3540 |
| **Fat** | 762 | 746.72 - 777.59 | 20 | Linescan (VNIR) | 0.2416 | 0.2447 | 0.2179 | 0.2280 |
| | | 741.84 - 780.83 | 4 | Linescan (NIR) | 0.2564 | 0.2619 | 0.2055 | 0.2353 |
| | | 740.67 - 780.73 | 4 | Snapshot (NIR) | 0.2564 | 0.2619 | 0.2399 | 0.2638 |
| | 830 | 814.95 - 844.20 | 19 | Linescan (VNIR) | 0.0793 | 0.0766 | 0.0769 | 0.0719 |
| | | 818.20 - 845.82 | 3 | Linescan (NIR) | 0.1219 | 0.1129 | 0.1205 | 0.0740 |
| | | 817.51 - 845.03 | 3 | Snapshot (NIR) | 0.1487 | 0.1280 | 0.1941 | 0.1470 |
| | 930 | 915.68 - 944.92 | 19 | Linescan (VNIR) | 0.1934 | 0.1825 | 0.2755 | 0.2203 |
| | | 915.68 - 941.67 | 4 | Linescan (NIR) | 0.0941 | 0.1154 | 0.1139 | 0.0210 |
| | | 914.48 - 940.95 | 4 | Snapshot (NIR) | 0.1819 | 0.1805 | 0.1965 | 0.1687 |
| **Water** | 976 | 949.80 - 995.29 | 25 | Linescan (VNIR) | 0.1082 | 0.1204 | 0.0749 | 0.1040 |

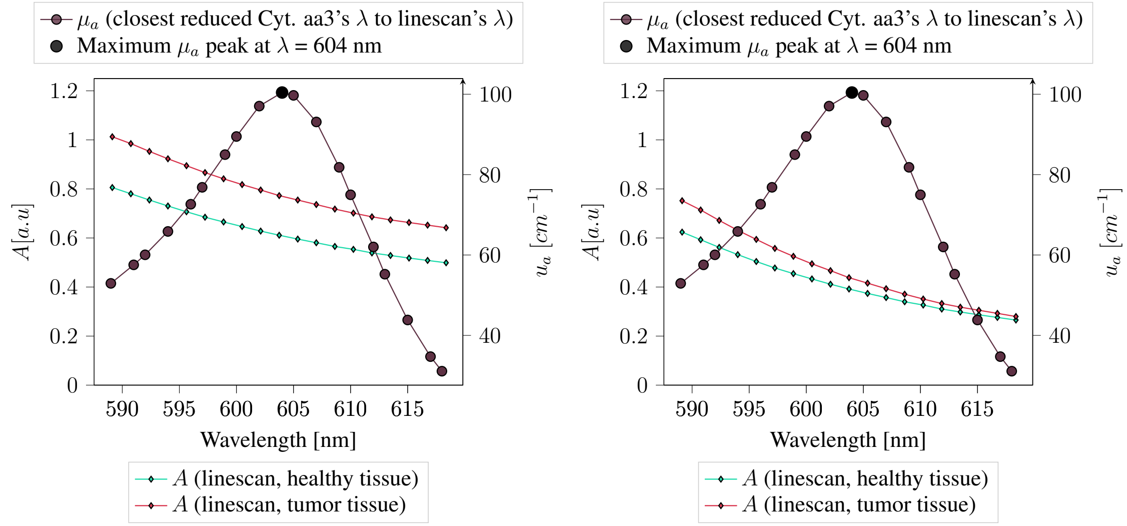


(a) Calibrated and denoised

(b) Calibrated, denoised, and normalized

**Fig S4** Absorption coefficients values, $\mu_a$, of Cyt. aa3 in its reduced state compared with the absorbance measurements, $A$, taken for the healthy and pathological tissues captured with the hyperspectral cameras.

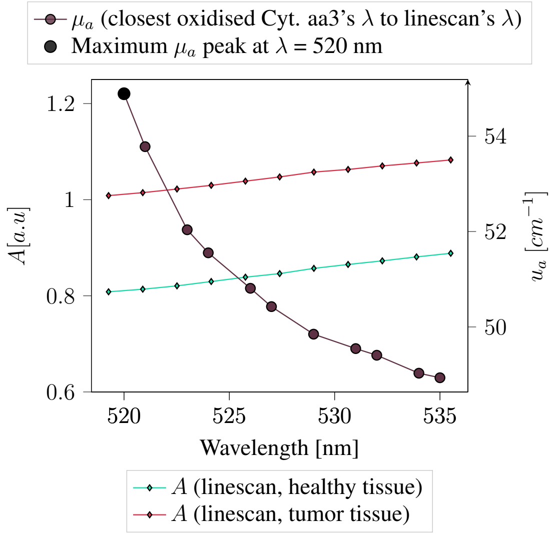


(a) Calibrated and denoised

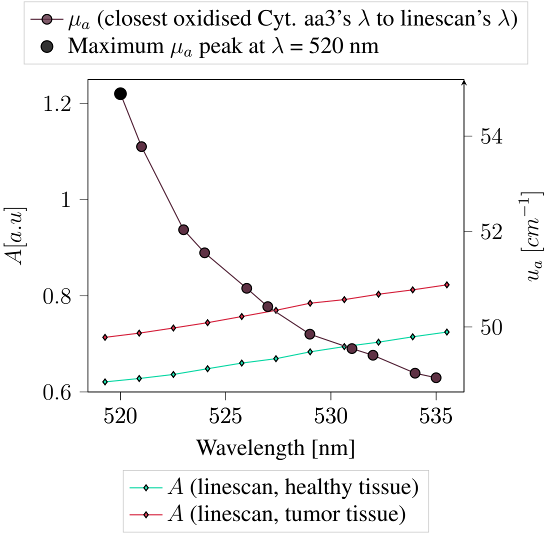


(b) Calibrated, denoised, and normalized

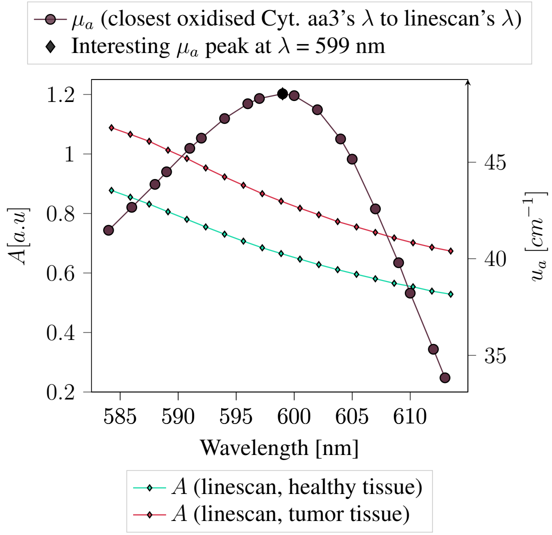


(c) Calibrated and denoised

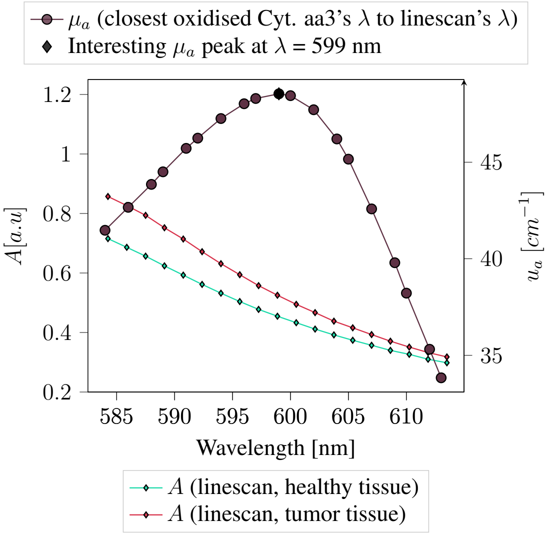


(d) Calibrated, denoised, and normalized

**Fig S5** Absorption coefficients values, $\mu_a$, of Cyt. aa3 in its oxidised state compared with the absorbance measurements, $A$, taken for the healthy and pathological tissues captured with the hyperspectral cameras.

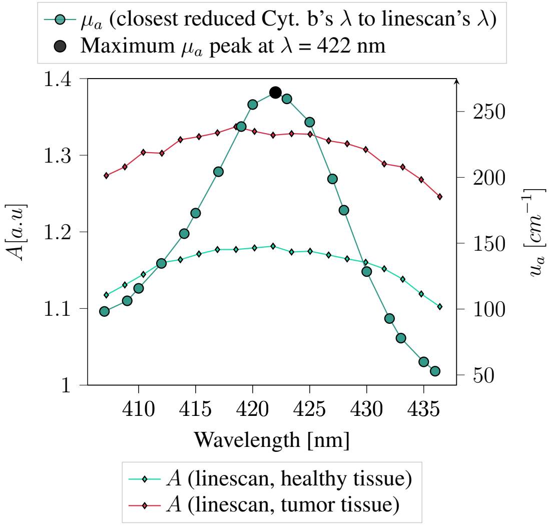


(a) Calibrated and denoised

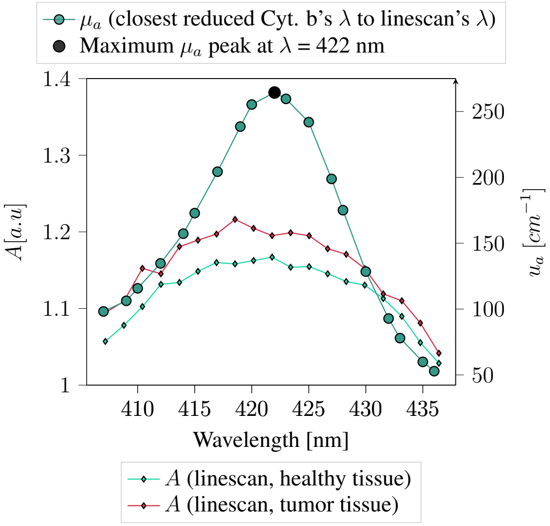


(b) Calibrated, denoised, and normalized

**Fig S6** Absorption coefficients values, $\mu_a$, of Cyt. b in its reduced state compared with the absorbance measurements, $A$, taken for the healthy and pathological tissues captured with the hyperspectral cameras.

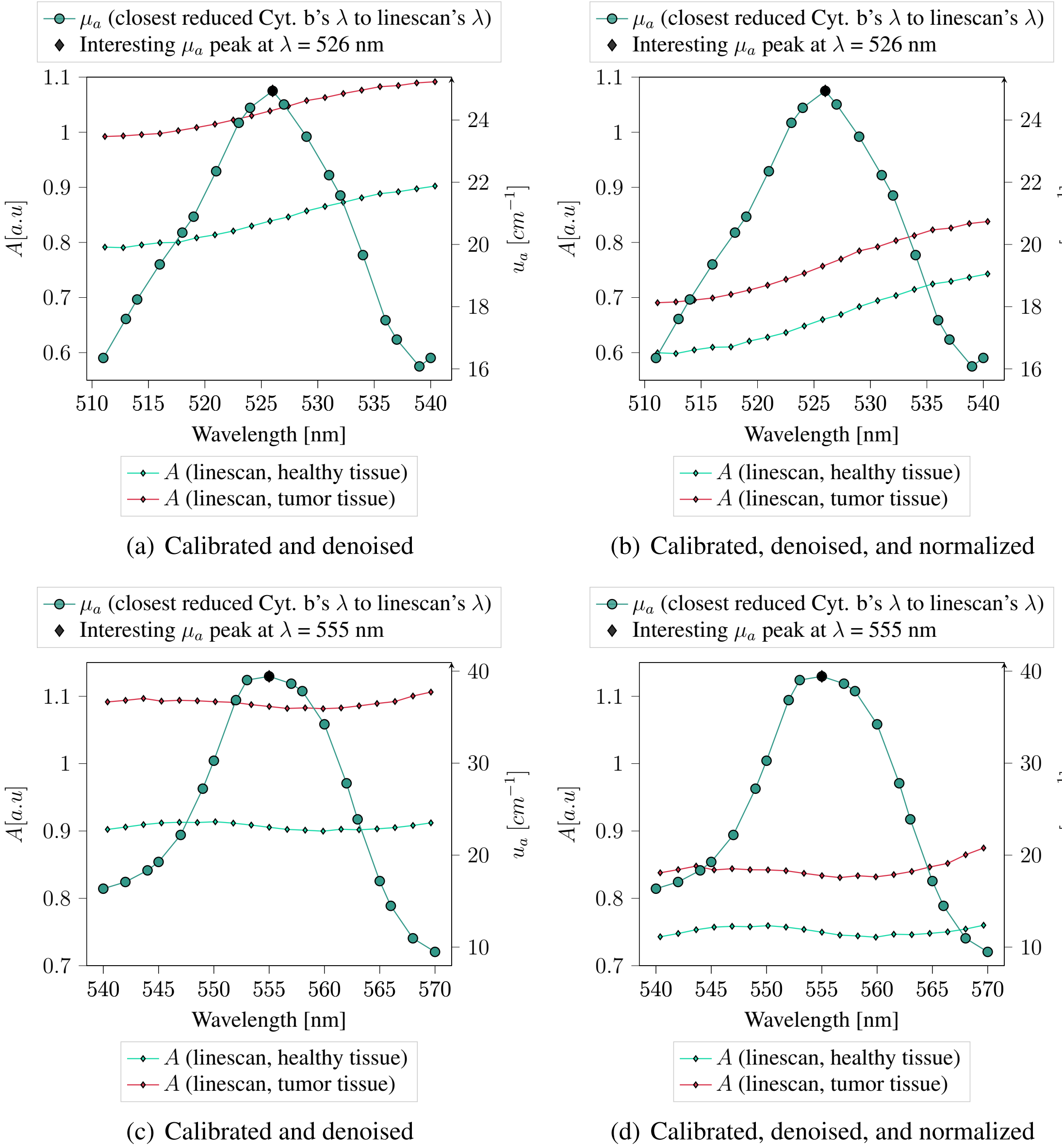


**Fig S7** Absorption coefficients values, $\mu_a$, of Cyt. b in its reduced state compared with the absorbance measurements, $A$, taken for the healthy and pathological tissues captured with the hyperspectral cameras.

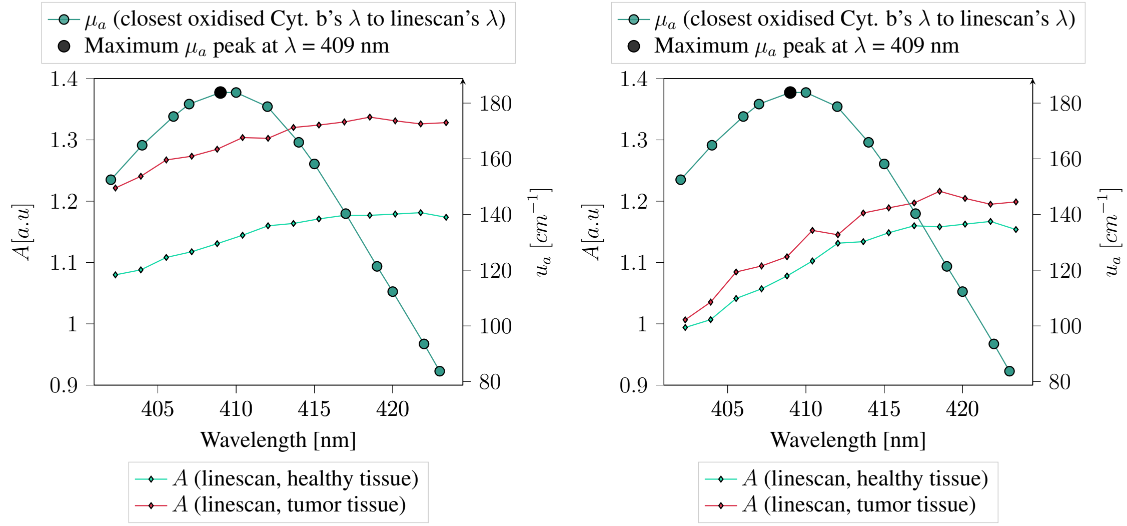


(a) Calibrated and denoised

(b) Calibrated, denoised, and normalized

**Fig S8** Absorption coefficients values, $\mu_a$, of Cyt. b in its oxidised state compared with the absorbance measurements, $A$, taken for the healthy and pathological tissues captured with the hyperspectral cameras.

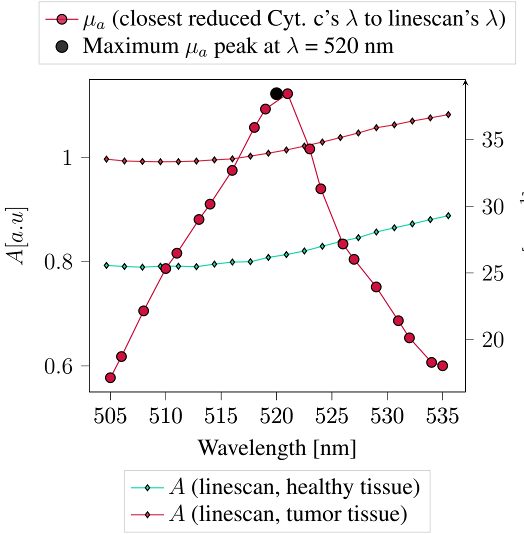


(a) Calibrated and denoised

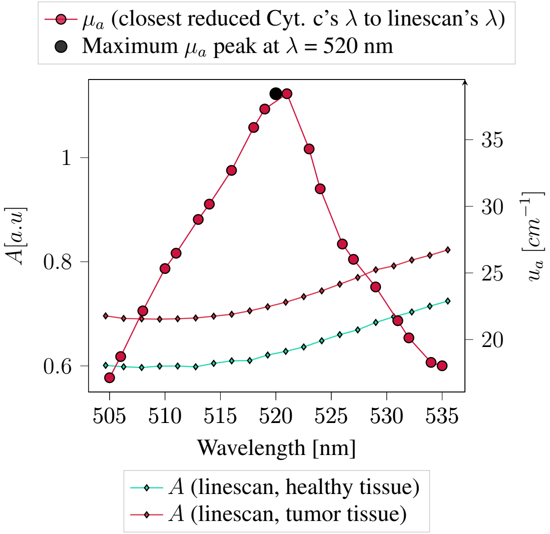


(b) Calibrated, denoised, and normalized

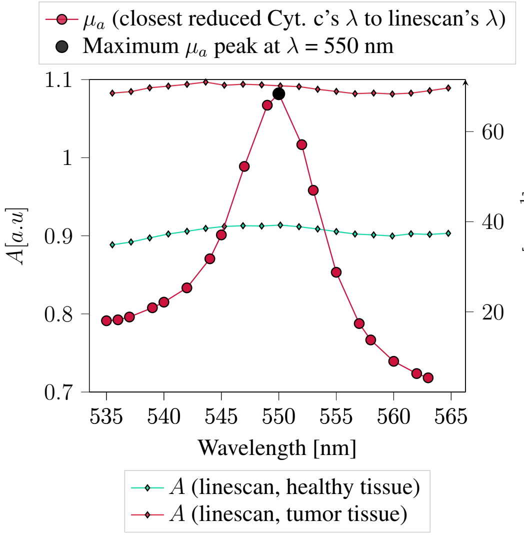


(c) Calibrated and denoised

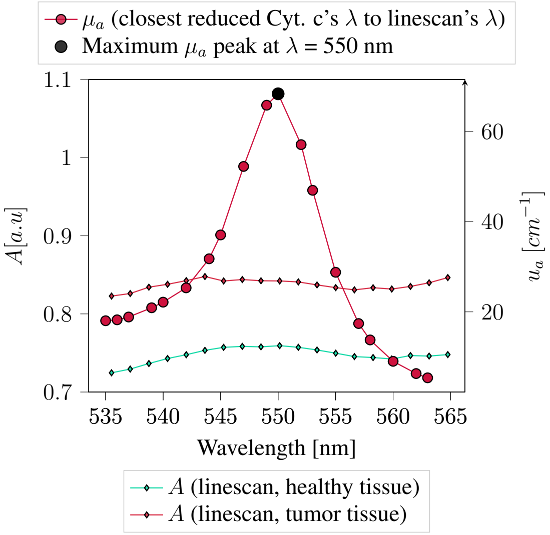


(d) Calibrated, denoised, and normalized

**Fig S9** Absorption coefficients values, $\mu_a$, of Cyt. c in its reduced state compared with the absorbance measurements, $A$, taken for the healthy and pathological tissues captured with the hyperspectral cameras.

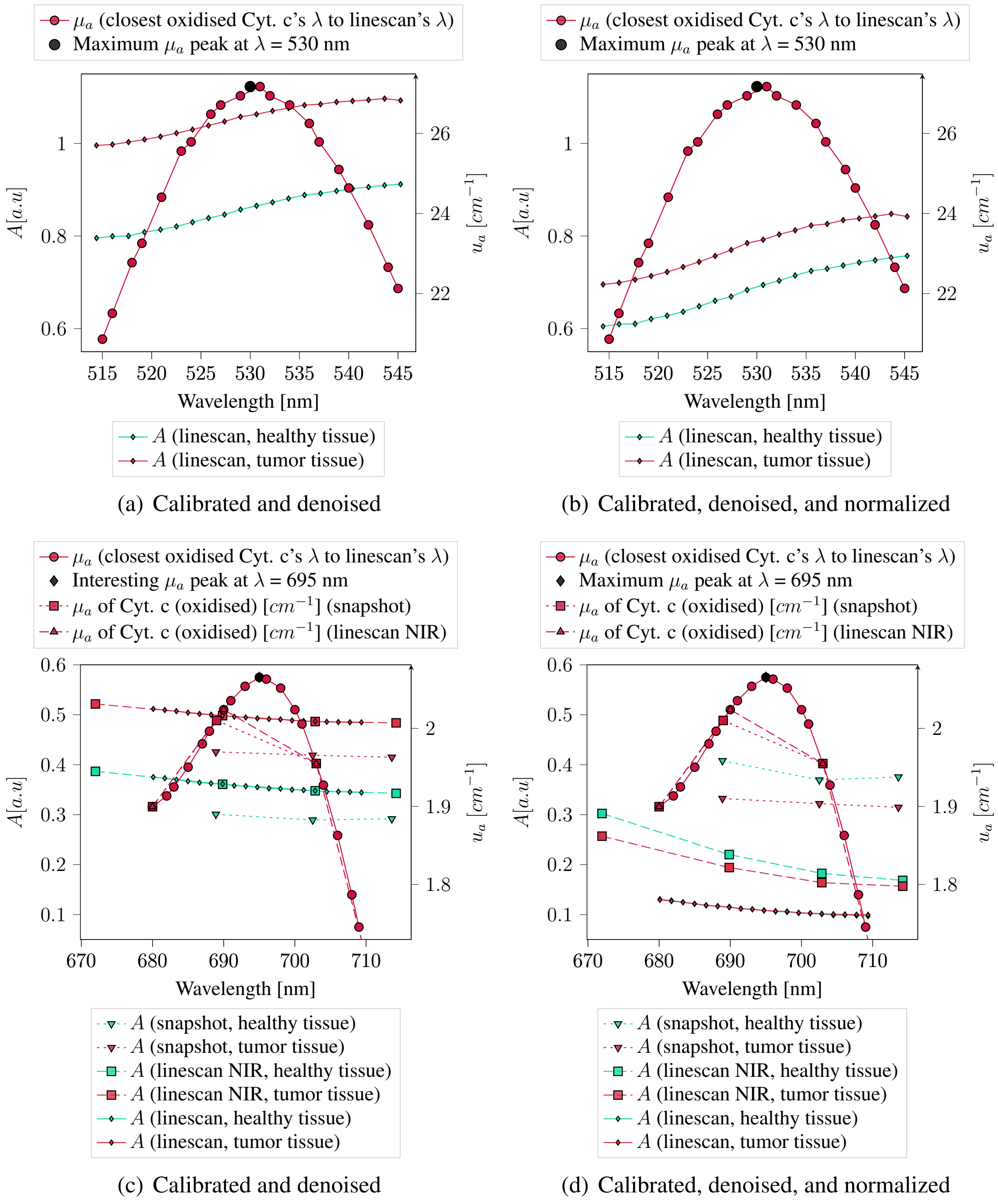


**Fig S10** Absorption coefficients values, $\mu_a$, of Cyt. c in its oxidised state compared with the absorbance measurements, $A$, taken for the healthy and pathological tissues captured with the hyperspectral cameras.

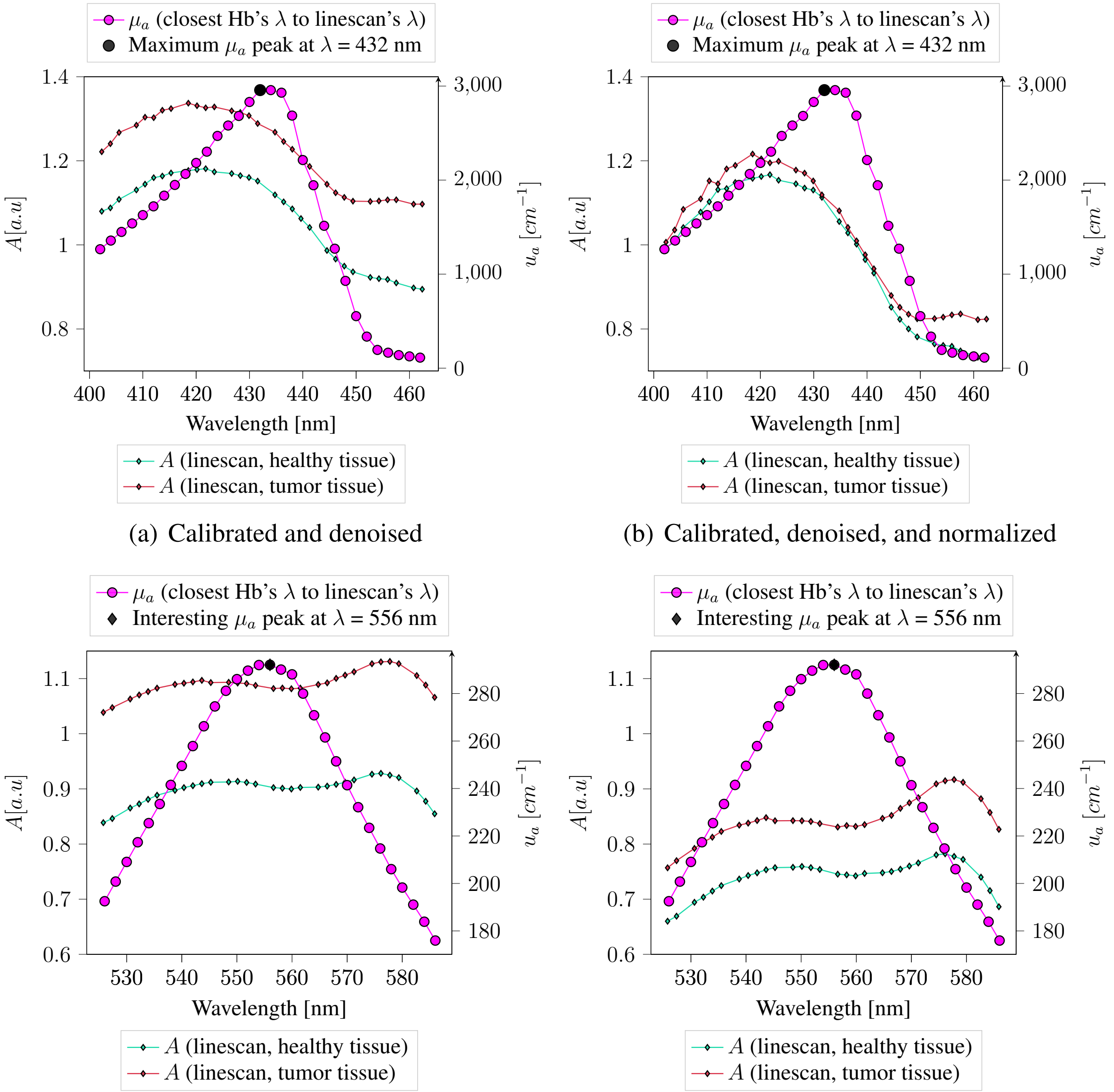


(a) Calibrated and denoised

(b) Calibrated, denoised, and normalized

(c) Calibrated and denoised

(d) Calibrated, denoised, and normalized

**Fig S11** Absorption coefficients values, $\mu_a$, of Hb compared with the absorbance measurements, $A$, taken for the healthy and pathological tissues captured with the hyperspectral cameras.

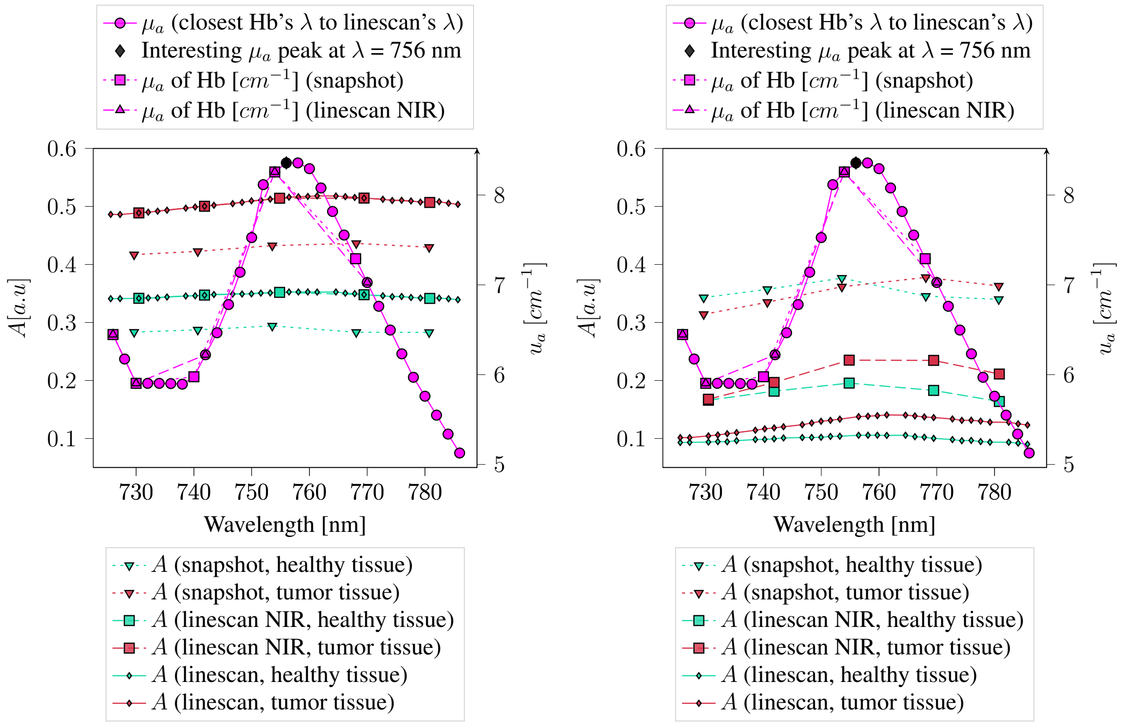


(a) Calibrated and denoised (b) Calibrated, denoised, and normalized

**Fig S12** Absorption coefficients values, $\mu_a$, of Hb compared with the absorbance measurements, $A$, taken for the healthy and pathological tissues captured with the hyperspectral cameras.

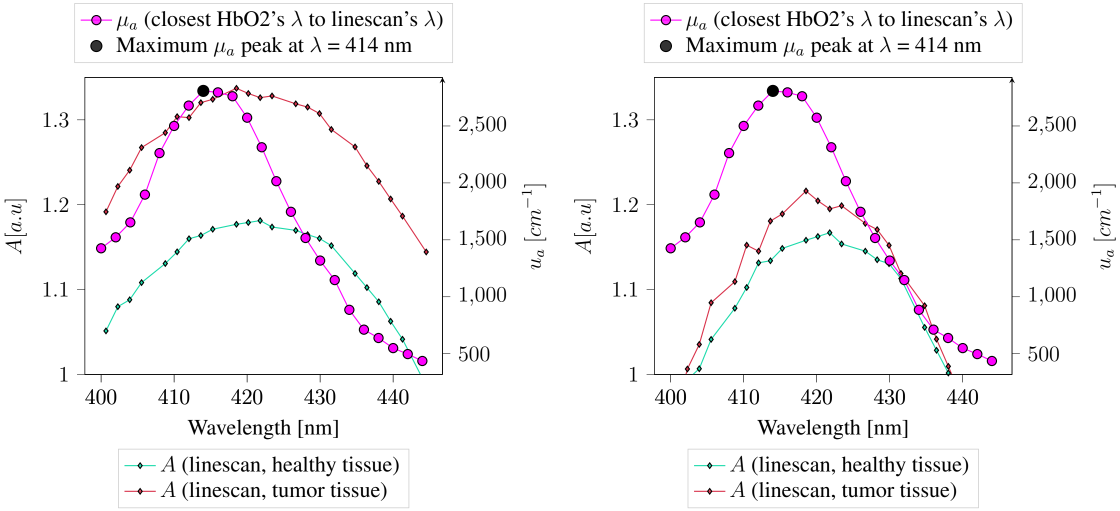


(a) Calibrated and denoised (b) Calibrated, denoised, and normalized

**Fig S13** Absorption coefficients values, $\mu_a$, of HbO2 compared with the absorbance measurements, $A$, taken for the healthy and pathological tissues captured with the hyperspectral cameras.

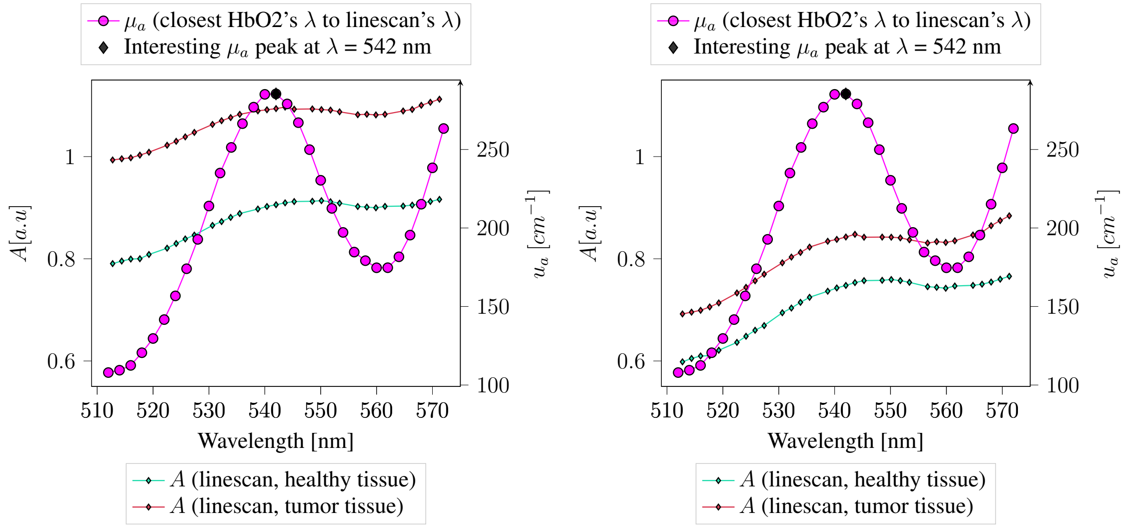


(a) Calibrated and denoised
(b) Calibrated, denoised, and normalized

**Fig S14** Absorption coefficients values, $\mu_a$, of HbO2 compared with the absorbance measurements, $A$, taken for the healthy and pathological tissues captured with the hyperspectral cameras.

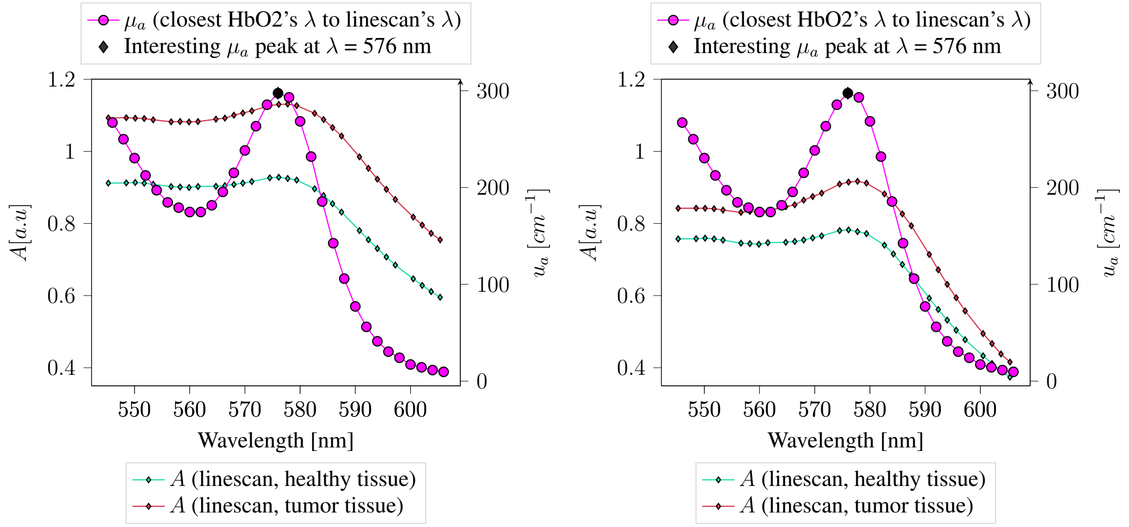


(a) Calibrated and denoised
(b) Calibrated, denoised, and normalized

**Fig S15** Absorption coefficients values, $\mu_a$, of HbO2 compared with the absorbance measurements, $A$, taken for the healthy and pathological tissues captured with the hyperspectral cameras.

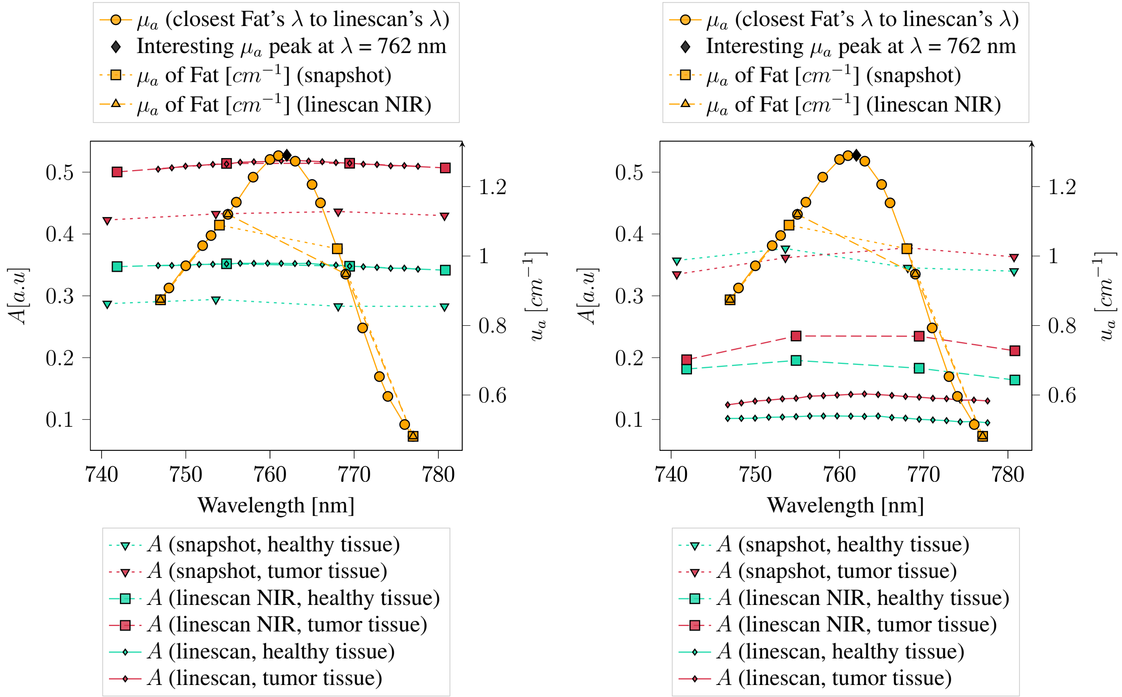


(a) Calibrated and denoised (b) Calibrated, denoised, and normalized

**Fig S16** Absorption coefficients values, $\mu_a$, of fat compared with the absorbance measurements, $A$, taken for the healthy and pathological tissues captured with the hyperspectral cameras.

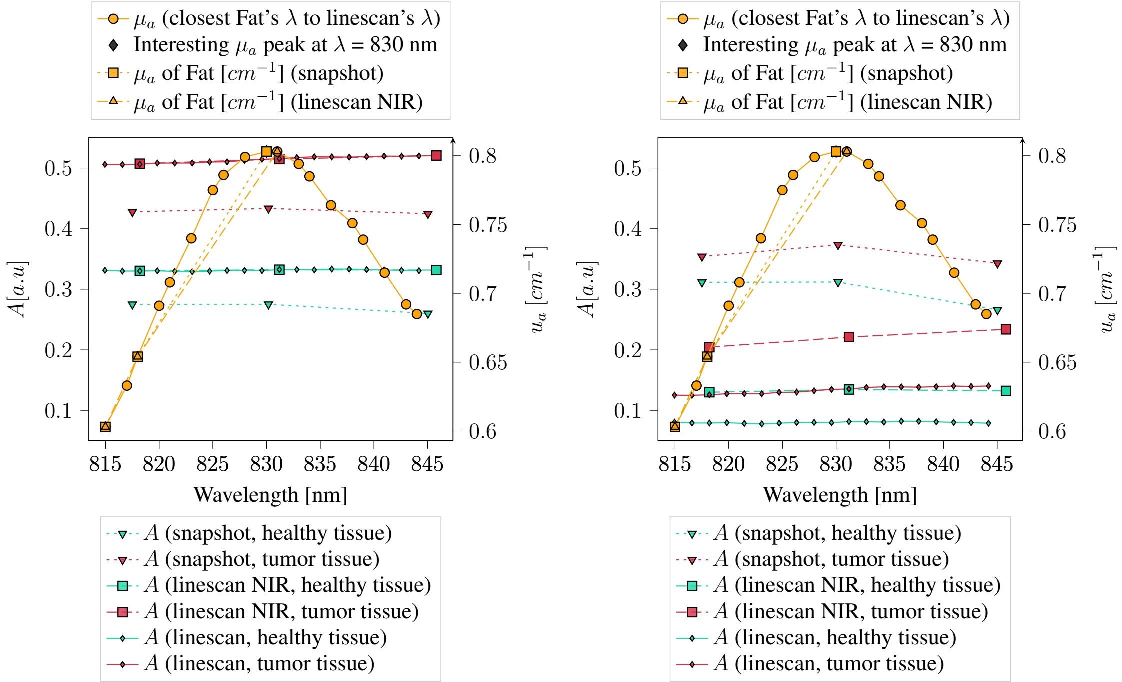


(a) Calibrated and denoised (b) Calibrated, denoised, and normalized

**Fig S17** Absorption coefficients values, $\mu_a$, of fat compared with the absorbance measurements, $A$, taken for the healthy and pathological tissues captured with the hyperspectral cameras.

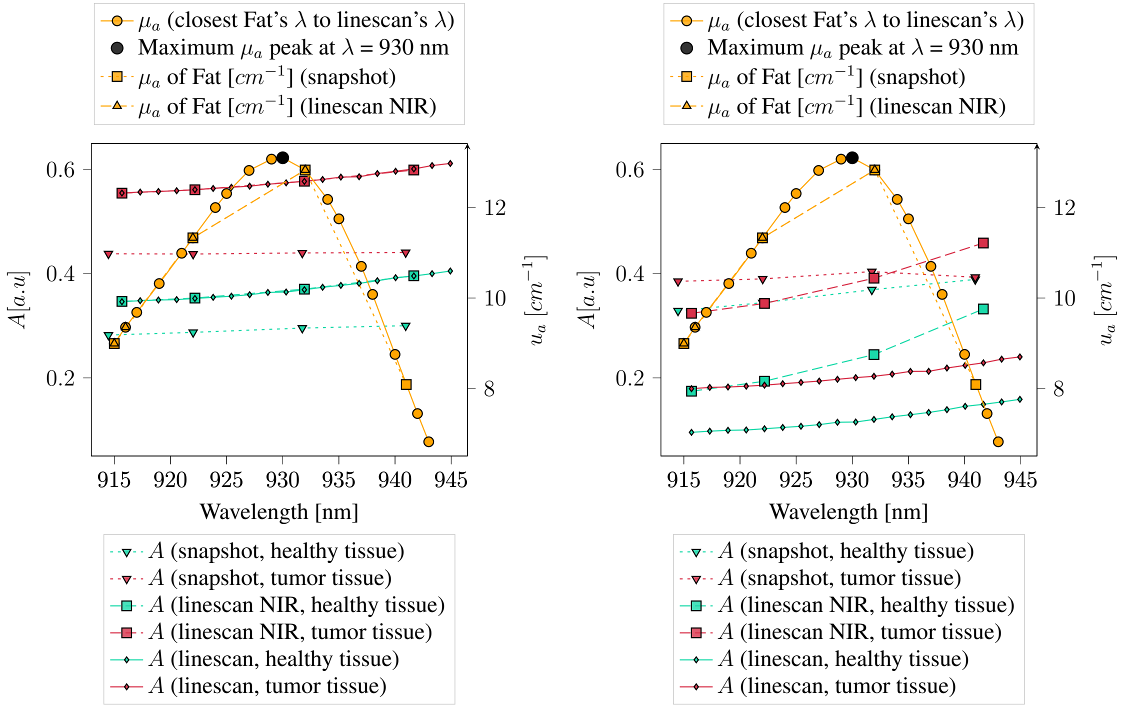


(a) Calibrated and denoised (b) Calibrated, denoised, and normalized

**Fig S18** Absorption coefficients values, $\mu_a$, of fat compared with the absorbance measurements, $A$, taken for the healthy and pathological tissues captured with the hyperspectral cameras.

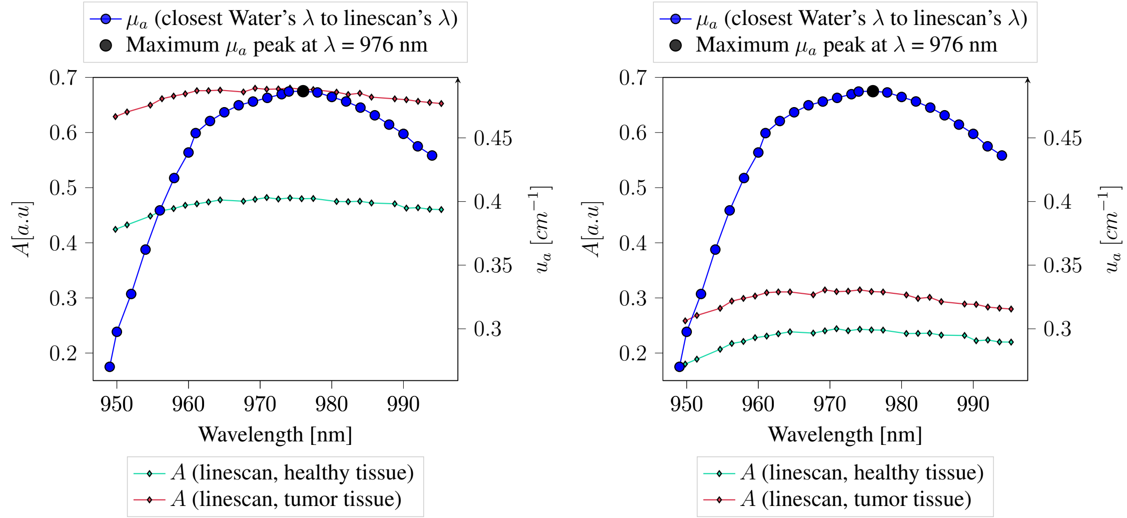


(a) Calibrated and denoised (b) Calibrated, denoised, and normalized

**Fig S19** Absorption coefficients values, $\mu_a$, of water compared with the absorbance measurements, $A$, taken for the healthy and pathological tissues captured with the hyperspectral cameras.

*References*


1 H. Fabelo, M. Halicek, S. Ortega, *et al.*, "Deep learning-based framework for in vivo identification of glioblastoma tumor using hyperspectral images of human brain," *Sensors* **19**(4) (2019).

2 R. Leon, S. Ortega, I. A. Cruz-Guerrero, *et al.*, "Hyperspectral imaging benchmark based on machine learning for intraoperative brain tumour detection," *npj Precision Oncology* **7**, 119 (2023).

3 H. Abramczyk, J. M. Surmacki, B. Brozek-Pluska, *et al.*, "Revision of commonly accepted warburg mechanism of cancer development: Redox-sensitive mitochondrial cytochromes in breast and brain cancers by raman imaging," *Cancers* **13**(11) (2021).

4 P. R. Bargo, S. A. Prahl, T. T. Goodell, *et al.*, "In vivo determination of optical properties of normal and tumor tissue with white light reflectance and an empirical light transport model during endoscopy," *Journal of biomedical optics* **10**(3), 034018–034018 (2005).

5 A. Cerussi, N. Shah, D. Hsiang, *et al.*, "In vivo absorption, scattering, and physiologic properties of 58 malignant breast tumors determined by broadband diffuse optical spectroscopy," *Journal of biomedical optics* **11**(4), 044005–044005 (2006).

6 F. Poulon, H. Mehidine, M. Juchaux, *et al.*, "Optical properties, spectral, and lifetime measurements of central nervous system tumors in humans," *Scientific reports* **7**(1), 13995 (2017).

7 R. Mühle, W. Markgraf, A. Hilsmann, *et al.*, "Comparison of different spectral cameras for image-guided organ transplantation," *Journal of Biomedical Optics* **26**(7), 076007–076007 (2021).
